# A multi-phenotype analysis reveals 19 novel susceptibility loci for basal cell carcinoma and 15 for squamous cell carcinoma

**DOI:** 10.1101/2022.03.06.22271725

**Authors:** Mathias Seviiri, Matthew H. Law, Jue-Sheng Ong, Puya Gharahkhani, Pierre Fontanillas, 23andMe Research Team, Catherine M. Olsen, David C. Whiteman, Stuart MacGregor

**Affiliations:** Statistical Genetics Lab, QIMR Berghofer Medical Research Institute, 300 Herston Road, Herston QLD 4006, Australia; School of Biomedical Sciences, Faculty of Health, and Center for Genomics and Personalised Health, Queensland University of Technology, 60 Musk Avenue, Kelvin Grove QLD 4059, Australia; 23andMe, Inc, Sunnyvale, CA, USA; Cancer Control Group, QIMR Berghofer Medical Research Institute, 300 Herston Road, Herston QLD 4006, Australia; Faculty of Medicine, University of Queensland, 20 Weightman St, Herston QLD 4006, Australia

## Abstract

Basal cell carcinoma (BCC) and squamous cell carcinoma (SCC) are the most common forms of skin cancer. There is genetic overlap between skin cancers, pigmentation traits, and autoimmune diseases. We use linkage disequilibrium score regression to identify 20 traits (melanoma, pigmentation traits, autoimmune diseases, and blood biochemistry biomarkers) with a high genetic correlation (*r*_g_ > 10%, P < 0.05) with BCC (20,791 cases and 286,893 controls in the UK Biobank) and SCC (7,402 cases and 286,892 controls in the UK Biobank), and use a multi-trait genetic analysis to identify 78 and 69 independent genome-wide significant (P < 5 × 10^-8^) susceptibility loci for BCC and SCC respectively; 19 BCC and 15 SCC loci are both novel and replicated (P < 0.05) in a large independent cohort; 23andMe, Inc (BCC: 251,963 cases and 2,271,667 controls, and SCC: 134,700 cases and 2,394,699 controls. Novel loci are implicated in BCC/SCC development and progression (e.g. *CDKL1*), pigmentation (e.g. *DSTYK*), cardiometabolic pathways (e.g. *FADS2*), and immune-regulatory pathways including; innate immunity against coronaviruses (e.g. *IFIH1*), and HIV-1 viral load modulation and disease progression (e.g. *CCR5*). We also report a powerful and optimised BCC polygenic risk score that enables effective risk stratification for keratinocyte cancer in a large prospective Canadian Longitudinal Study of Aging (794 cases and 18139 controls); e.g. percentage of participants reclassified; MTAG_PRS_ = 36.57%, 95% CI = 35.89-37.26% versus UKB_PRS_= 33.23%, 95% CI=32.56-33.91%).

## INTRODUCTION

Keratinocyte cancers (KC), including basal cell carcinoma (BCC) and squamous cell carcinoma (SCC), are the most commonly diagnosed cancers globally. KC resulted in over 5.4 million diagnoses and $8 billion dollars in expenditure in the US in 2011 alone ^1^, while in Australia they account for > 24% of all cancer diagnoses ^2^, and impose a huge economic burden on the health sector costing over AUD $700 million for treatment annually ^3^. KC is responsible for upto 8,700 deaths a year in the US ^4^. The relative rates, and morbidity, from KC is even higher in Australia ^5^. BCC and SCC share many common risk factors including sun exposure, skin and hair pigmentation, and immunosuppression.

Skin cancers, and pigmentation traits and autoimmune diseases have several susceptibility genes overlapping ^6–9^. For example, several variants in pigmentation genes *ASIP/RALY, IRF4, MC1R, OCA2, SLC45A2* and *TYR*, are associated with BCC, SCC and melanoma ^8,10^. Shared immune regulatory genes in the *HLA* and *LPP* regions have been found to influence susceptibility to BCC, SCC, melanoma and autoimmune diseases such as rheumatoid arthritis, vitiligo, type 1 diabetes and psoriasis ^6–9^. There are also some tumor-genesis related genes which are expressed in both KC and other non-skin cancers. For example, oncogene *TNS3* which is overregulated in BCC is also associated with breast cancer, lung cancer and prostate cancer ^8,11,12^. Furthermore, *HAL* at 12q23.1 has been found to be associated with KC risk ^13^ as well as vitamin D levels ^14^. However, standard single GWAS meta-analysis approaches are unable to utilise this multi-trait genetic overlap to further explore the genetic risk for BCC, and SCC.

Multivariate GWAS approaches, such as multi-trait analysis of GWAS (MTAG) ^15^, can draw on this overlapping genetics to identify new risk regions (here for BCC or SCC). MTAG is a generalisation of inverse-variance-weighted meta-analysis that importantly accounts for incomplete genetic correlation, and sample overlap, between GWAS. A key property of MTAG is that it outputs estimates of trait-specific effect sizes and p-values for each of the input traits - in this case BCC or SCC. We have previously used MTAG to identify loci for KC based on the genetic correlation between BCC and SCC only ^13^. BCC and SCC are different in terms of polygenicity and aetiology and therefore, we sought to identify novel susceptibility genetic loci for BCC and SCC by exploring their genetic overlap with melanoma, pigmentation traits, autoimmune diseases, and blood biochemistry biomarkers in a multi-phenotype analysis of GWAS.

## RESULTS

### Genetic correlation

Using linkage disequilibrium score (LDSC) regression ^16^, 20 phenotypes were significantly genetically correlated (*P* < 0.05, *rg* > 10%) with either BCC or SCC (**Figure 1** and **Supplementary Table 1**). In the first instance, 35 phenotypes that we considered as possibly correlated with skin cancer (including body mass index) were excluded for not meeting aforementioned criteria above (**Supplementary Table 2**). Using the same selection criteria, no additional new phenotypes were included following analysis using collated GWAS summary statistics (over 700 phenotypes) in the LD hub database ^17^. In total, subsequent analyses included 22 genetically correlated traits; cancers; BCC and SCC GWAS from the UK Biobank (UKB) ^18,19^, a cutaneous melanoma GWAS meta-analysis ^20^, KC from the QSkin Sun and Health Study (QSkin) ^21^, KC from the Electronic Medical Records and Genomics Network (eMERGE) cohort ^22,23^ and all-cancer from the Resource for Genetic Epidemiology Research on Aging (GERA) cohort ^24^; skin and hair pigmentation related traits; skin burn type (QSkin), red hair (QSkin), hair colour excluding red hair (UKB), skin colour (UKB), and mole count excluding melanoma cases (QSkin), autoimmune conditions; type 1 diabetes and hypothyroidism ^25^, and vitiligo ^26^, lifestyle-related traits; educational attainment in years spent in school ^27^ and smoking (cigarettes per day) ^28^, and biochemistry blood biomarkers from the UKB; aspartate aminotransferase, C-reactive protein, albumin, and gamma-glutamyl transferase, glucose and vitamin D (adjusted for monthly variation). The sample sizes and phenotype measurement for all the included and excluded traits are presented in **Supplementary Tables 3** and **Supplementary Table 2** respectively.

**Figure 1:**
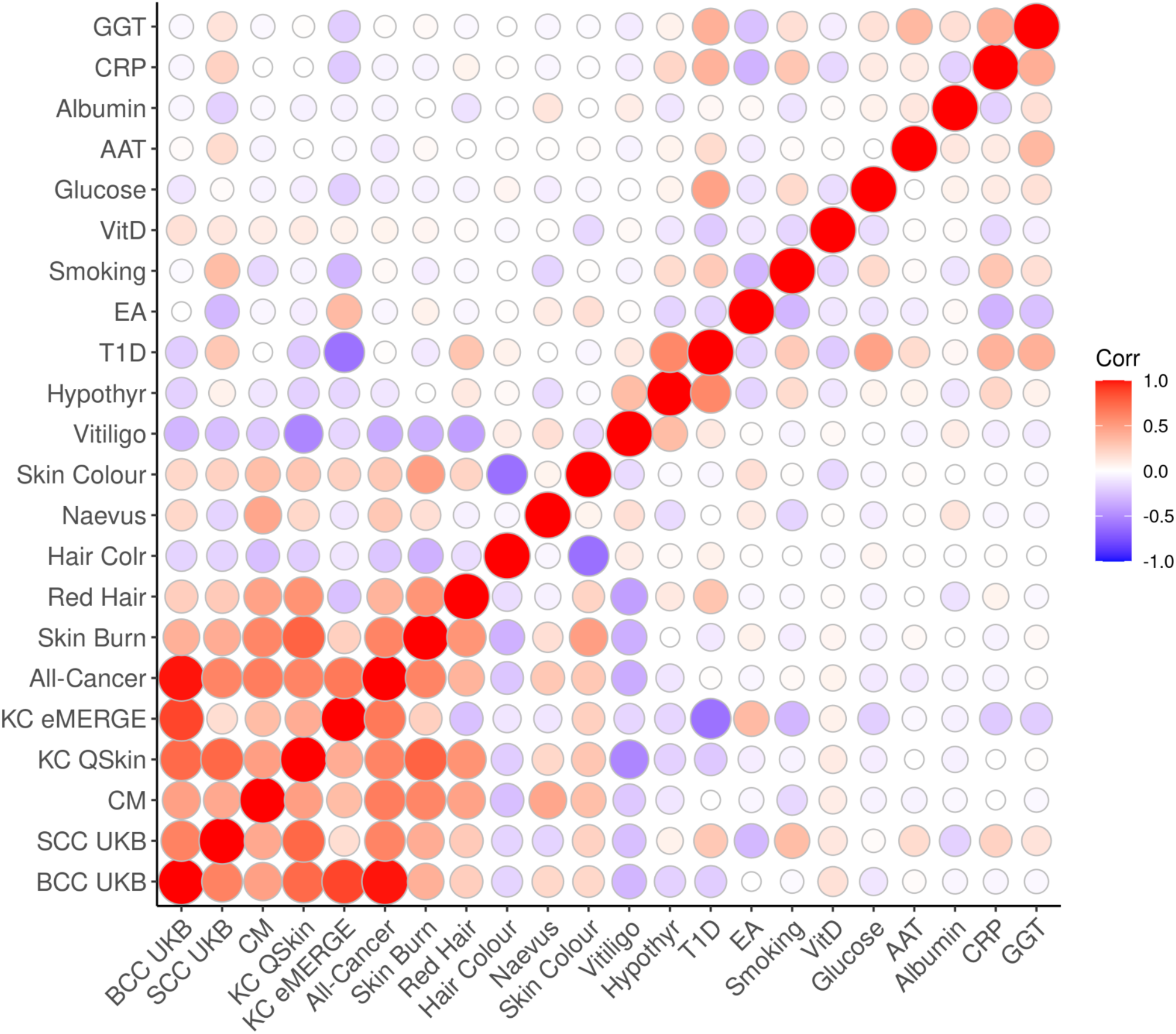
Heatmap for the genetic correlation between 22 traits with a significant correlation with either BCC or SCC. Bivariate genetic correlation 22 traits that were significantly correlated (P<0.05, rg >10%) with the UKB BCC or SCC GWAS. Abbreviations: basal cell carcinoma in the UK Biobank (BCC UKB), squamous cell carcinoma in the UK Biobank (SCC UKB), cutaneous melanoma (CM), keratinocyte cancer in the QSkin cohort (KC QSkin), keratinocyte cancer in the eMERGE cohort (KC eMERGE), hypothyroidism (Hypothyr), type 1 diabetes (T1D), education attainment (EA), vitamin D (VitD), aspartate aminotransferase (AAT), C-reactive protein (CRP), gamma-glutamyl transferase (GGT) and correlation (Corr).

### Discovery of genome-wide significant susceptibility loci for BCC and SCC

Adding 20 traits genetically correlated with either BCC or SCC (r_g_ > 0.1, *P* < 0.05) (from UKB) increased the effective sample sizes for BCC and SCC by 2.6 and 8.3 times respectively. Using the MTAG approach we identified 78 and 69 independent genome-wide significant (P < 5 × 10^-8^) susceptibility loci for BCC (**Figure 2** and **Supplementary Table 4**) and SCC (**Figure 3** and **Supplementary Table 5**) respectively. Although the results for the peak single nucleotide polymorphisms (SNPs) were more significant following the MTAG analysis due to the greater statistical power, the log (odds ratio) effect sizes for the MTAG output and the respective UKB BCC or SCC GWAS inputs were highly concordant. For BCC the Pearson’s correlation of effect sizes was 0.93 (95% confidence interval [CI]= 0.89-0.96, *P* < 2.20 x 10^-16^; **Figure 4a**). Similarly, concordance was high for SCC loci (Pearson’s correlation = 0.71, 95% CI=0.57-0.81, P = 7.34 x 10^-12^; **Figure 4b**).

**Figure 2:**
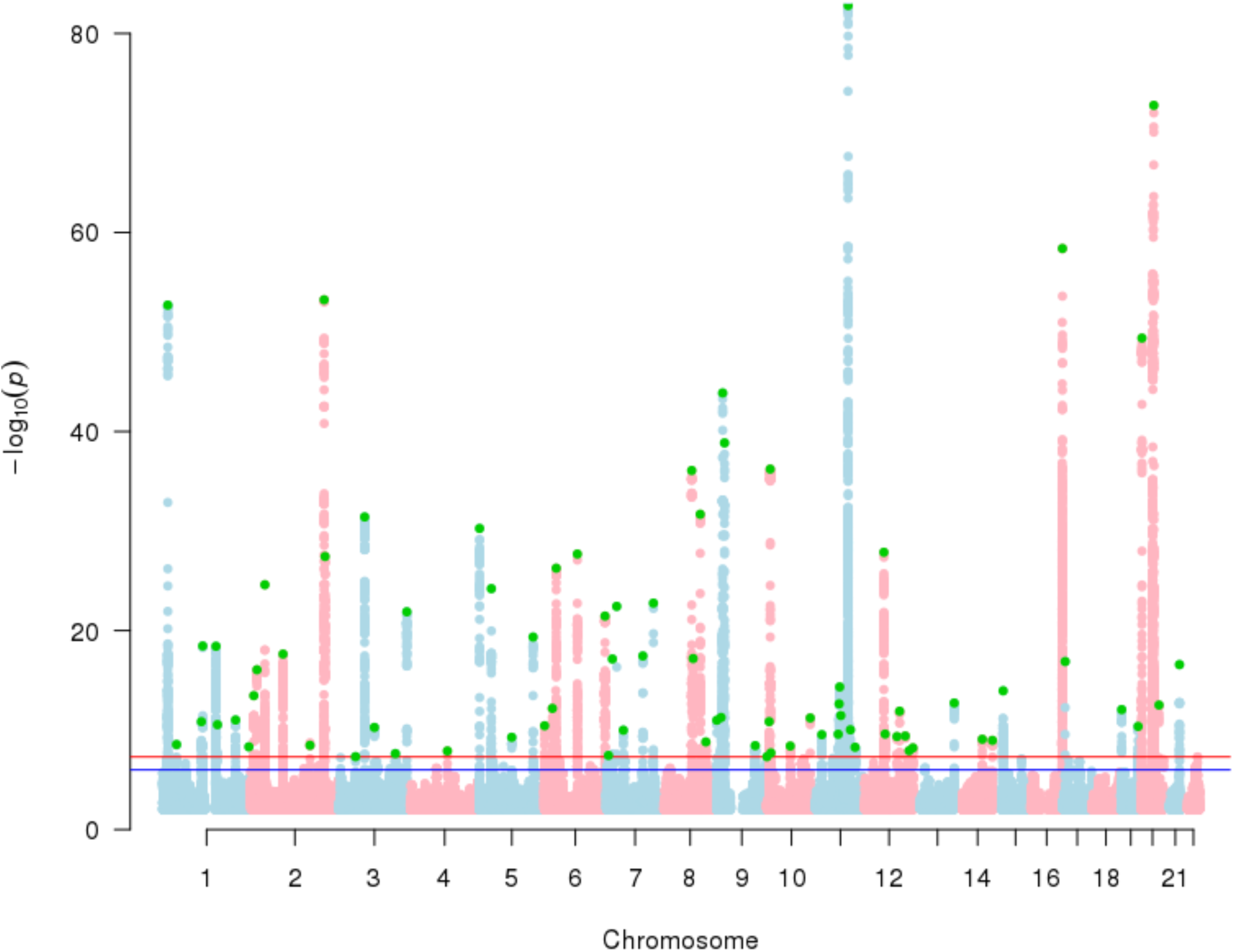
Manhattan plot for basal cell carcinoma susceptibility. The Manhattan plot shows the association between SNPs and basal cell carcinoma susceptibility based on the MTAG approach. The Y-axis represents the level of significance recorded in negative log 10 (P-value), whilst the X-axis represents the chromosome 1-22, alternated with light blue and light pink colours. The horizontal blue line represents a suggestive level of significance at P-value =10^-6^, while the red one represents the genome-wide level of significance; P = 5x 10^-8^. The green dots represent the 78 genome-wide significant independent loci for basal cell carcinoma susceptibility.

**Figure 3:**
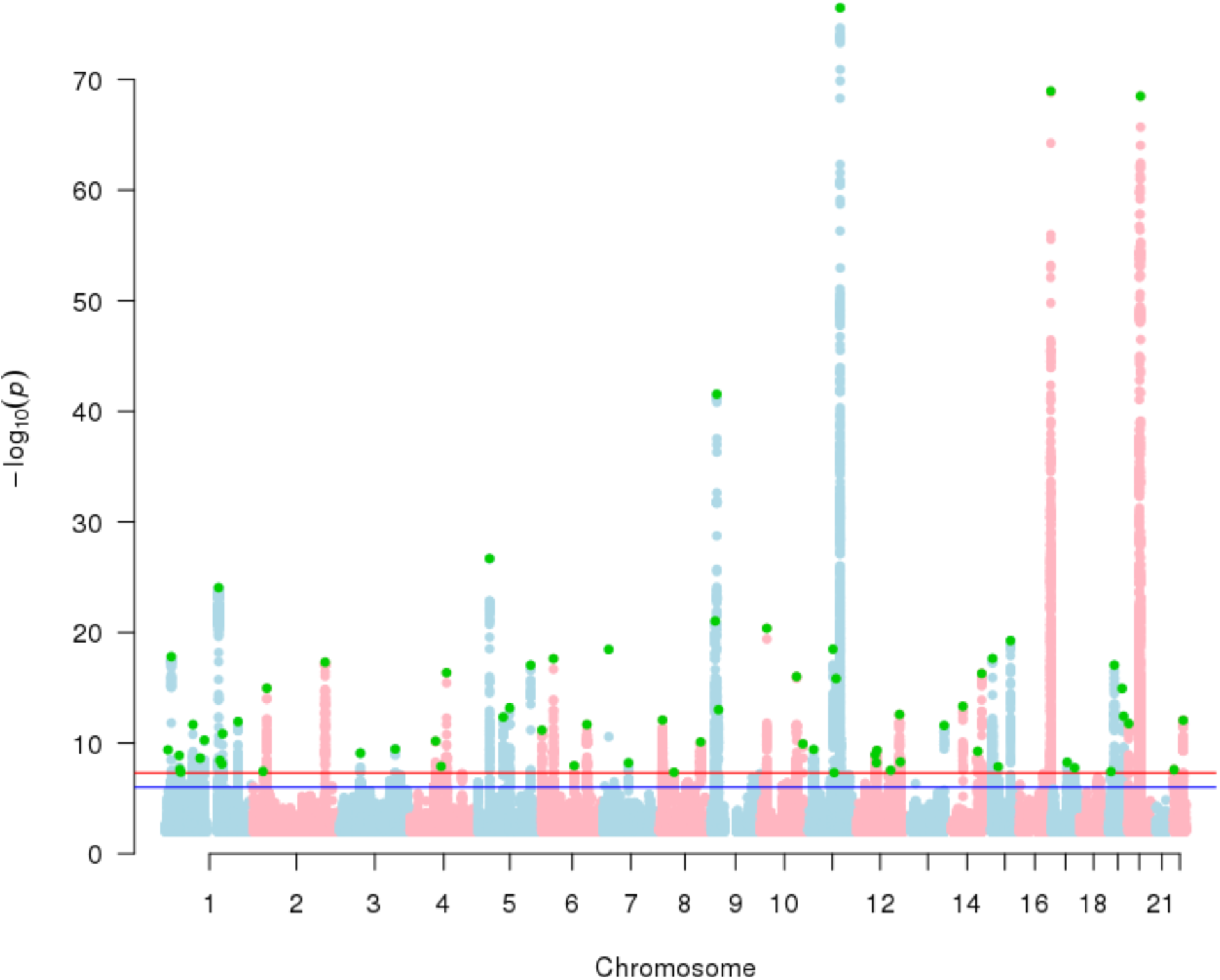
Manhattan plot for squamous cell carcinoma susceptibility. The Manhattan plot shows the association between SNPs and squamous cell carcinoma susceptibility based on the MTAG approach. The Y-axis represents the level of significance recorded in negative log 10 (P-value), whilst the X-axis represents the chromosome 1-22, alternated with light blue and light pink colours. The horizontal blue line represents a suggestive level of significance at P-value =10^-6^, while the red one represents the genome-wide level of significance; P = 5x 10^-8^. The green dots represent the 69 genome-wide significant independent loci for squamous cell carcinoma susceptibility.

**Figure 4:**
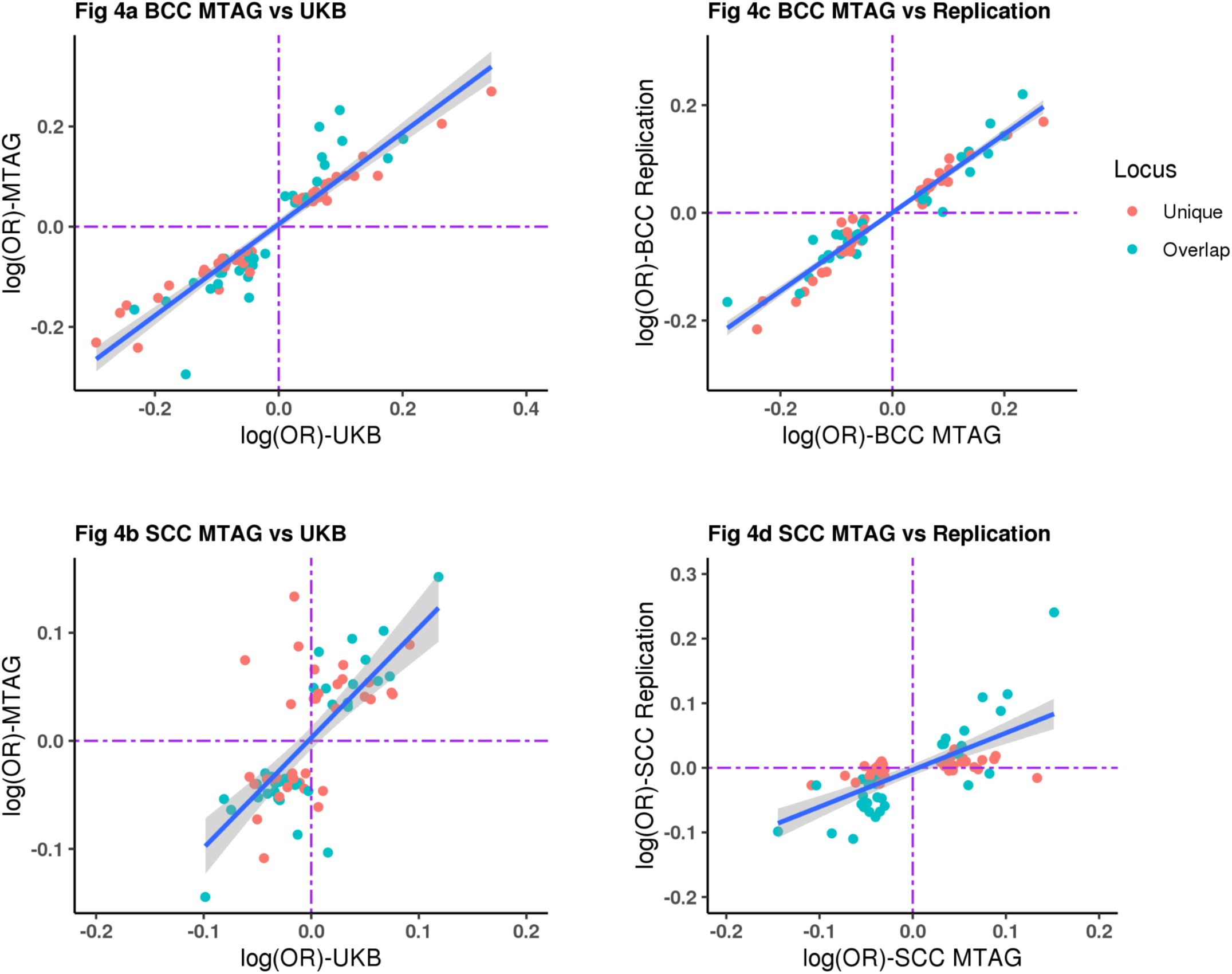
Concordance of the log (OR) effect estimates for MTAG versus UK single trait GWAS and 23andMe replication. Figure 4 shows the comparison of the effect estimates in log (odds ratio) for both basal cell carcinoma and squamous cell carcinoma based on the respective MTAG approach results versus UKB single trait GWAS and replication results from 23andMe. The blue line is the line of best fit with the 95% confidence intervals. The blue dots represent loci that overlap between BCC and SCC, whilst the red dots show the loci that are respectively unique to BCC or SCC. The dotted purple lines represent null effects (i.e. log (OR) =0). The Y- and X-axes represent log (OR). **4 a**: Shows BCC MTAG versus UKB BCC effect estimates, yielding a high concordance with a Pearson’s correlation of 0.93 (95% confidence interval [CI]= 0.89-0.96). **4 b**: Shows SCC MTAG versus UKB SCC effect estimates, yielding a high concordance; Pearson’s correlation = 0.71, 95% CI=0.57-0.81). **4 c**: Shows BCC MTAG versus BCC replication (23andMe) effect estimates, yielding a high concordance i.e. Pearson’s correlation = 0.97 (95% CI = 0.95-0.98). **4 d**: Shows SCC MTAG versus SCC replication effect estimates, also resulting in a high correlation i.e. 0.69 (95% CI=0.55-0.80).

In the 23andMe, Inc replication sample (252,931 cases and 2,281,246 controls), 71 of the 78 susceptibility loci for BCC replicated at genome-wide level (*P* < 5 x 10^-8^), 74 replicated after Bonferroni correction (*P* = 6.49 x 10^-4^), and 77 loci replicated at a nominal *P* = 0.05 (**Supplementary Table 4**). There was high concordance with the BCC effect estimates between the MTAG and the replication set with Pearson’s correlation = 0.97 (95% CI = 0.95-0.98, *P* = 2.20 x 10^-16^; **Figure 4c**). Of the 69 susceptibility loci for SCC, 25 replicated at genome-wide level (*P* = 5 x 10^-8^), 31 replicated after Bonferroni correction (*P* = 7.24 x 10^-4^) and 38 loci replicated at a nominal *P* = 0.05 in the 23andMe cohort (135,214 cases and 2,404,735 controls) (**Supplementary Table 5**). For SCC there was also high concordance with the effect estimates between the MTAG and the replication set with Pearson’s correlation = 0.69 (95% CI=0.55-0.80, P = 3.48 x 10^-11^; **Figure 4d**).

### Description of the BCC and SCC novel loci

A locus was considered novel for BCC or SCC if it had not previously been significantly associated with either BCC, SCC or KC at genome-wide level (*P* <5 x 10^-8^), and if it replicated at minimum P < 0.05) in the 23andMe replication cohort. By this criterion we identified 19 and 15 novel loci for BCC (**Table 1**) and SCC (**Table 2**), respectively. After correcting for multiple testing, 18 and 12 novel loci replicated for BCC (Bonferroni P < 2.38 x 10^-3^) **(Supplementary Tables 4**) and SCC (Bonferroni P < 2.63 x 10^-3^) **(Supplementary Tables 5**) respectively. The novel loci were annotated to the pigmentation, cardiometabolic and immune-regulatory pathways, whilst others are known loci for cutaneous melanoma susceptibility. We now discuss the novel loci in broader biological groups. For loci that are unique to BCC or SCC, or overlap between BCC and SCC refer to **Table 1** and **Table 2**.

**Table 1:**
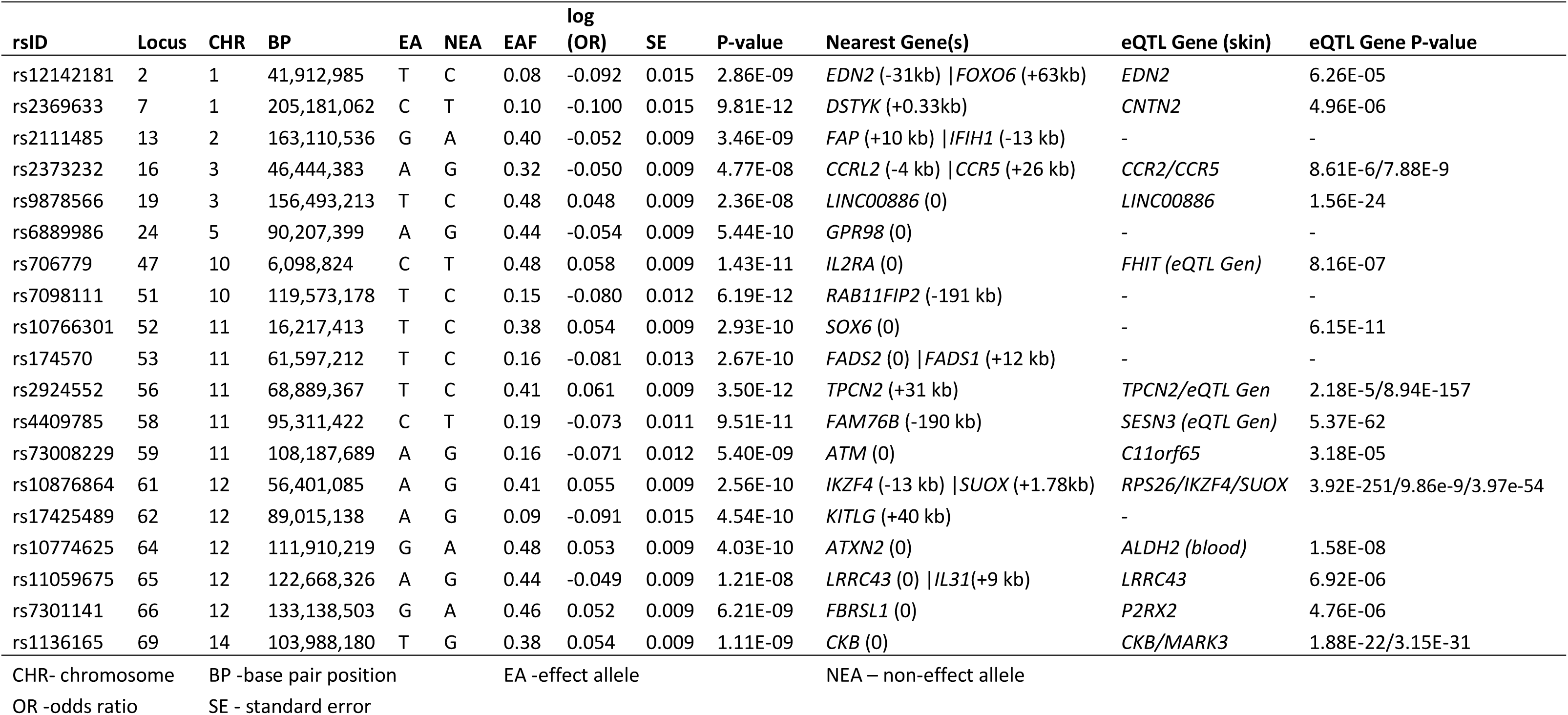
BCC susceptibility novel loci that replicated at P < 0.05 in 23andMe cohort.

**Table 2:** SCC susceptibility novel loci that replicated at P < 0.05 in 23andMe cohort

| rsID | Locus | CHR | BP | EA | NEA | EAF | log (OR) | SE | P | Nearest Gene | eQTL Gene (skin) | eQTL Gene P-value |
| --- | --- | --- | --- | --- | --- | --- | --- | --- | --- | --- | --- | --- |
| rs3768321 | 3 | 1 | 40,035,928 | T | G | 0.19 | 0.041 | 0.007 | 1.32E-09 | PABPC4 (0) | PABPC4 | 2.65E-28 |
| rs12142181 | 4 | 1 | 41,912,985 | T | C | 0.08 | -0.054 | 0.010 | 2.41E-08 | EDN2 (-31.46kb) | EDN2 | 6.26E-05 |
| rs17391694 | 6 | 1 | 78,623,626 | T | C | 0.10 | 0.054 | 0.008 | 2.12E-12 | GIPC2 (+20.51kb) | FUBP1 | 5.49E-15 |
| rs3851290 | 13 | 1 | 205,149,508 | T | C | 0.39 | -0.039 | 0.006 | 1.17E-12 | DSTYK (0) | DSTYK | 5.52E-18 |
| rs1260326 | 14 | 2 | 27,730,940 | C | T | 0.41 | -0.030 | 0.005 | 3.57E-08 | GCKR (0) | NRBP1 | 3.09E-18 |
| rs9878566 | 18 | 3 | 156,493,213 | T | C | 0.48 | 0.034 | 0.005 | 3.52E-10 | LINC00886 (0) | LINC00886 | 1.56E-24 |
| rs6889986 | 24 | 5 | 90,207,399 | A | G | 0.44 | -0.041 | 0.005 | 6.75E-14 | GPR98 (0) | - | 8.94E-11 |
| rs77758638 | 33 | 8 | 42,014,917 | T | C | 0.11 | 0.045 | 0.008 | 4.35E-08 | AP3M2 (0) | POLB/AP3M2 | 4.39E-26 |
| rs35563099 | 40 | 10 | 119,572,403 | T | C | 0.15 | -0.046 | 0.007 | 1.19E-10 | RAB11FIP2 (-192kb) | - |  |
| rs2924552 | 42 | 11 | 68,889,367 | T | C | 0.41 | 0.049 | 0.005 | 3.19E-19 | TPCN2 (+31.3kb) | TPCN2 | 2.18E-05 |
| rs10899466 | 44 | 11 | 78,013,674 | A | G | 0.18 | -0.061 | 0.007 | 1.47E-16 | GAB2 (0) | GAB2 | 1.24E-13 |
| rs10876864 | 47 | 12 | 56,401,085 | A | G | 0.41 | 0.031 | 0.005 | 5.95E-09 | SUOX (+1.78kb) \| IKZF4 (-13 kb) | SUOX/IKZF4 | 3.97E-54/9.86E-9 |
| rs142004400 | 53 | 14 | 50,829,560 | C | A | 0.03 | -0.109 | 0.014 | 4.83E-14 | CDKL1 (0) | CDKL1 (eQTL Gen) | 1.13E-19 |
| rs10141120 | 55 | 14 | 103,923,008 | C | T | 0.35 | -0.047 | 0.006 | 5.18E-17 | MARK3 (0) | MARK3/CKB | 7.60E-39/3.33E-31 |
| rs472385 | 57 | 15 | 44,186,844 | A | G | 0.25 | -0.035 | 0.006 | 1.37E-08 | FRMD5 (0) | AC011330.5 | 5.74E-19 |
| CHR- chromosome |  | BP -base pair position |  |  |  | EA -effect allele |  |  |  | NEA – non-effect allele |  |  |
| OR -odds ratio |  | SE - standard error |  |  |  |  |  |  |  |  |  |  |

### Novel loci for keratinocyte cancer development and progression

We identified seven novel loci with a potential role in the development and progression of keratinocyte cancer. rs10141120 in *MARK3*; *MARK3* is a cell cycle regulator involved in the DNA damage response (e.g. following radiotherapy or treatment with alkylating agents) ^29^, and implicated in carcinogenesis e.g. for hepatocellular carcinoma ^30^. In addition, rs10141120 is in LD with rs3825566 (r^2^=0.94) and rs55859054 (r^2^=0.96) which are lead SNPs for hair colour ^31,32^.

We also identified two novel variants for BCC and SCC; rs35563099 and rs7098111 respectively, near *RAB11FIP2* that were in high LD (r^2^=0.98). *RAB11FIP2* is overexpressed in colorectal and gastric cancer cells where it facilitates their migration leading to cancer metastases ^33,34^. It is likely that *RAB11FIP2* promotes keratinocyte cancer progression. rs35563099 is a lead SNP for skin low tanning response ^35^ and in LD with rs11198112 (r^2^=0.99) for sunburns^31^. rs7098111 also is in LD with SNPs for skin/hair colour (rs11198112, r^2^=0.57), freckles (rs10444039, r^2^=0.99) and sunburns (rs11198112, r^2^=0.98)^31,36,37^.

rs10899466 in *GAB2* is in LD with rs10899501 for hair colour (r^2^=0.80) ^31,38^. Following inflammatory stimuli (e.g. by cytokines) *GAB2* is required in inflammatory signalling during tumorigenesis ^39,40^. It enhances cancer cell proliferation e.g. in breast cancer ^41,42^. Another novel variant was rs10766301, an intronic variant in *SOX6*. Although the direct role of *SOX6* in keratinocyte cancer biology is unknown, it facilitates apoptosis in colorectal cancer, esophageal squamous cell carcinoma, pancreatic and ovarian cancer ^43–46^. Conversely, its downregulation facilitates cancer progression ^43,45,47^. rs10766301 is in LD with rs2953060 (r^2^=0.60) for sunburns ^31^.

rs142004400 in *CDKL1* promotes tumor cell proliferation, migration and invasion in melanoma, colorectal cancer, and breast cancer ^48–50^, whose downregulation facilitates apoptosis ^48^. We also identified rs472385 in *FRMD5; FRMD5* modulates tumour progression by regulating cancer cell mobility and ROCK1-triggered kinase activity ^51,52^. Its expression is downregulated in renal, breast and colorectal cancers ^52^. rs472385 is also in LD with rs35654783 (r^2^=0.76), a lead SNP for diastolic blood pressure ^53^.

We also identified rs10876864 near *SUOX* (+1.78kb); expression of *SUOX* is associated with both proliferation and progression of oral squamous cell carcinoma, gastric cancer and hepatocellular carcinoma ^54–56^. In addition, eQTL analysis showed that rs10876864 was strongly linked to expression of *SUOX* and *RPS26* in skin tissues (**Table 1** and **Table 2**). *RPS26* regulates the tumour suppression activities of *p53* in response to DNA damage ^57^. Thus, it is possible that rs10876864 is involved in proliferation and progression of keratinocyte cancers. However, it is also likely that rs10876864 might have pleiotropic effects on KC through immunomodulating pathways since it is also near *IKZF4* (−13.6kb); *IKZF4* is required to suppress/maintain FOXP3+ regulatory T cells ^58,59^, important in auto-immunity and self-recognition. It is in LD with lead SNPs for auto-immune traits e.g. T1D, and allergic disease, rheumatoid arthritis (in LD with rs773125, r^2^=0.83) ^60–62^, immune-suppressive medication use e.g. glucocorticoids (in LD with rs1689510, r^2^=0.72), thyroid preparations (in LD with rs7302200, r^2^= 0.71) and anti-asthmatic adrenergics inhalant use (in LD with rs34415530, r^2^=0.72) ^63^.

### Novel loci in the immune regulation pathway

Previous studies have reported a relationship between immune response and skin cancer and a number of our novel loci for BCC and SCC suggested links to immune regulatory processes. rs2373232 is an intergenic variant between *CCRL2* (−4.337kb) and *CCR5* (+26.69kb) in high linkage disequilibrium (LD) r^2^=1, with rs1015164 (in *CCR5*), a known SNP for HIV-1 viral load variation ^64,65^. *CCR5* is generally involved in coordination of the immune response ^66^, and specifically regulates HIV-1 viral load and progression ^67,68^. *CCRL2* is involved in regulating immune responses induced by chemokines ^69^.

rs2111485 is an intergenic variant between *FAP* (+10.49kb) and *IFIH1* (−13.05kb) and in high LD (r^2^=0.89) with a nonsense SNP rs1990760 in *IFIH1; IFIH1* is involved in innate (anti-viral) immune response (e.g. against coronaviruses), autoimmunity and autoinflammatory response ^70–72^. In addition, *FAP* plays a pro-tumourigenic role in several cancers including breast, colorectal, gastric and esophageal cancer ^73^, and also facilitates immunosuppression to enhance colorectal and gastric cancer progression ^74,75^.

rs10774625, an intronic variant in *ATXN2,* is a lead SNP for hypothyroidism susceptibility ^76^, and in high LD (r^2^=0.9) with a missense SNP rs3184504 in *SH2B3*; *SH2B3* is involved in mediating immune cell stimulation and inflammatory signalling ^77,78^. Another novel SNP rs4409785 near *FAM76B* ( -190.7kb) is an *eQTL* for *SESN3* (in *eQTLGEN*), which regulates senescence in T-cells to influence the immune response during aging ^79,80^. rs4409785 is also associated with thyroid preparations (medication use), hypothyroidism, vitiligo and rheumatoid arthritis ^31,63,81,82^. At 10p15.1 rs706779 is an intronic variant in the *interleukin 2 receptor subunit alpha* (*IL2RA)* gene and lies a strong transcriptionally active enhancer in immune cells ^83^. *IL2RA* controls the immune response (tolerance) by modulating the function of regulatory T-cells ^84^. rs706779 is a lead SNP for T1D, Crohn’s disease, vitiligo and in LD with rs7090530 (r^2^=0.73) for hypothyroidism and thyroid medication ^26,31,60,63,85^. rs11059675 in *LRRC43*, and near *IL31* (+9.58kb) is a lead SNP for psoriasis ^86^, and is in LD with rs7968808 (r^2^= 0.98), a lead SNP for eczema ^31^. *IL31* induces and modulates skin allergic diseases ^87^. rs17391694 near *GIPC2* (+20.51kb) is a lead SNP for Crohn’s disease, an inflammatory autoimmune disorder ^85^, and lung cancer ^88^.

### Cardiometabolic pathway

For both BCC and SCC, we also identified novel loci that have been previously reported as being associated with cardiometabolic biomarkers. The BCC novel loci for this pathway included; rs174570 in *FADS2*, rs10774625 in *ATXN2*, and rs1136165 near *CKB* in BMI, whilst for SCC they included; rs3768321 in *PABPC4*, and rs1260326 near *GCKR*.

rs174570 is an intronic variant in *FADS2* and 12.68kb away from *FADS1* and it is a lead SNP for higher LDL cholesterol, total cholesterol and triglycerides levels, and is in LD with lead SNPs for PUFA levels in people of European descent e.g. rs174547 (r^2^=0.36), rs174577 (r^2^=0.34), and rs174538 (r^2^=0.42) ^89,90^. *FADS1/2* genes are involved in the downstream metabolism of the plasma omega-6 and omega-3 PUFA resulting in oncogenic inflammatory biomarkers (prostaglandins E, thromboxane A2, and leukotriene B) ^91^. rs1260326 in *GCKR* is a lead SNP for triglycerides, total cholesterol and fasting plasma glucose ^92,93^. Mutations in *GCKR* are known to be diabetogenic ^94^. rs3768321 in *PABPC4* is a lead SNP for HDL cholesterol ^92^.

Some novel variants in the cardiometabolic pathway had pleiotropic effects with pigmentation and autoimmune traits. For example rs1136165 in *CKB*, a lead SNP for BMI ^31^ is in LD with rs55859054 (r^2^=0.89) and rs3825566 (r^2^=0.88) which are associated with hair colour ^31,32^. rs10774625 in *ATXN2* is linked to a spectrum of cardiometabolic markers; diastolic and systolic blood pressure, CVD, coronary artery disease, and LDL cholesterol **(Supplementary Table 4**) ^31,53,95,96^, is also a lead SNP for immune regulatory phenotypes; hypothyroidism and T1D ^76,97^.

### Pigmentation pathways

Pigmentation is a crucial pathway in the development of BCC, SCC and melanoma. Known pigmentation genes like *MC1R* and *IRF4* play an important role in the genetic susceptibility to skin cancers and many new loci were associated with pigmentation traits. rs2924552 near *TPCN2* (+31.3kb); rare mutations in *TPCN2* results in blond rather than brown hair among Icelanders and the Dutch ^98^. rs9878566 in *LINC00886* is in perfect LD (r^2^=1.00) with rs9818780 for sunburns ^31^. rs6889986 in *GPR98* is in LD with lead SNPS for hair colour; rs60325490 (r^2^=0.69) and rs6860111 (r^2^=0.69) ^31,32^. rs77758638 in *AP3M2* is in LD with rs113060680 (r^2^=0.93) for hair colour and skin tanning response ^32^. However, there were also a number of loci with a potential role in KC initiation and progression that had pleiotropic effects with pigmentation traits (as explained above) e.g. rs35563099 and rs7098111 near *RAB11FIP2*.

### Novel BCC and SCC loci previously known for cutaneous melanoma susceptibility

Some novel BCC loci were previously known for CM; rs73008229 near *ATM* in LD (r^2^ ∼1) with rs1801516 a missense variant (i.e. L76I) in *ATM*; *ATM* has a cell cycle function in DNA damage response due to radiotoxicity after radiotherapy ^99–101^. rs1801516 has previously been associated with cutaneous melanoma ^102103,104^. Several other novel BCC SNPs are in high LD with lead SNPs for cutaneous melanoma; rs2369633 near *DSTYK* is in LD with rs11240396 (r^2^=0.73), rs6889986 near *GPR98* locus is in LD with rs10942621 (r^2^=0.71), rs10766301 in *SOX6* is in LD with rs2054095 (r^2^=0.65) ^20^.

### Unknown for any trait at genome-wide significance level

Some novel loci have not been associated with any trait before (at a genome-wide significance level); rs12142181 (near *RNA5SP45*), rs7301141 (near *FBRSL1*) for BCC and rs12142181 (near *RNA5SP45*) for SCC. However, rs12142181 is in LD with rs111599055 (r^2^=0.53) for early onset of prostate cancer ^105^.

### Gene-set pathways

After multiple correction testing (P = 0.05/18,188 genes; 2.75 × 10^-6^) gene set analysis revealed curated and gene ontology (GO) pathways that are important in development of keratinocyte cancer (**Supplementary Table 6**). A number of pathways are involved in melanogenesis (e.g. melanin biosynthesis, melanin biosynthetic process and melanosome membrane); a process which influences the nature of pigmentation traits and response to UV exposure. Genes in the “response to trabectedin” pathway are likely to play an important role in DNA damage response. Trabectedin is an alkylating agent used to treat certain cancers resulting in DNA damage. Other pathways are important in the downregulation of the immune response (e.g. “GO negative regulation of regulatory t cell differentiation”), and enhancement of the immune response (IL2-PI3K pathway, MHC class II receptor activity, and nuclear factor of activated T-cells (NFAT) pathway for development and function of regulatory T cells).

### BCC MTAG-derived polygenic risk score for KC prediction in the Canadian Longitudinal Study on Aging (CLSA)

During the validation of the PRSs, S5 (i.e. P < 10^-4^ with 273 SNPs for the MTAG_PRS_ and 462 SNPs for the UKB_PRS_) was the optimal PRS models for both MTAG_PRS_ and UKB_PRS_ with Nagelkerke R^2^ of 10.65% and 9.55% respectively (**Figure 5a**). The SNPs for the optimal models are presented in **Supplementary Table 7** and **Supplementary Table 8** for the UKB_PRS_ and MTAG_PRS_ respectively. When we tested the performance for both the UKB_PRS_ and MTAG_PRS_ in the CLSA (N=18,933), the MTAG_PRS_ outperformed the UKB_PRS_ in terms of association with KC risk, KC risk prediction, and stratification. For example, after adjusting for age at recruitment, sex and the first 10 PCs, the MTAG_PRS_ outperformed the UKB_PRS_ for association with KC risk i.e. MTAG_PRS_ OR = 1.66 95% CI = 1.55-1.79, P = 1.95×10^-41^ versus UKB_PRS_ OR = 1.56 95% CI = 1.45-1.67, *P* = 3.38×10^-33^ (**Figure 5b**). In addition, the net reclassification index for KC risk was greater for MTAG_PRS_ than the UKB_PRS_ (**Figure 5c**), when added to the base model containing age, sex and 10 PCs. Consequently, the MTAG_PRS_ compared to the UKB_PRS_ reclassified more participants for KC risk to the appropriate risk group (low risk, moderate risk and high risk) (i.e. percentage of people reclassified; MTAG_PRS_= 36.57% 95% CI = 35.89-37.26% versus UKB_PRS_= 33.23% 95% CI=32.56-33.91%) (**Figure 5d**).

**Figure 5:**
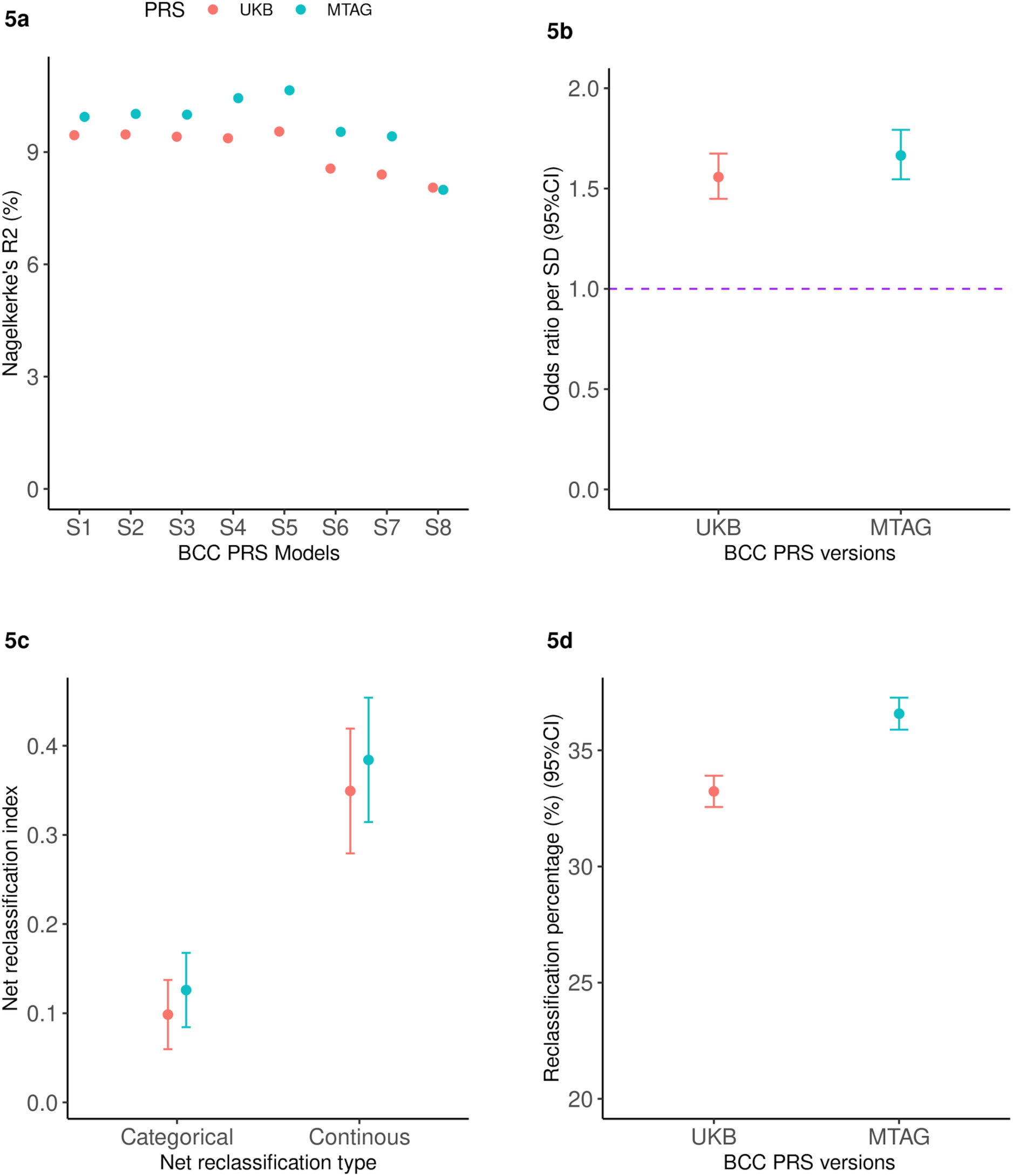
Validation and application of the basal cell carcinoma MTAG_PRS_ and UKB_PRS_ in the Canadian Longitudinal Study on Aging. The red colour represents the UKB PRS version whilst cyan indicates the MTAG-derived PRS. **5a:** Validation of the BCC MTAG_PRS_ and UKB_PRS_ models to select the best performing index based on clumped SNPs at S1 (P < 5×10^-8^), S2 (P < 10^-7^), S3 (P < 10^-6^), S4 (P < 10^-5^), S5 (P < 10^-4^), S6 (P < 10^-3^), S7 (P < 10^-2^) and S8 (P < 10^-1^) on the x-axis. The y-axis represents Nagelkerke’s R^2^ (%), a measure for model fitness. PRS model S1 and S5 are the optimal PRS models for UKB_PRS_ and MTAG_PRS_ respectively. **5b:** Shows and compares the association between the UKB_PRS_ and MTAG_PRS_ and KC risk in CLSA (N=18,515) expressed in odds ratios per standard deviation (y-axis) increase in the PRS adjusted for age, sex and the ancestral 10 PCs. **5c:** Illustrates that the MTAG_PRS_ performs better than the UKB_PRS_ based on both the categorical and continuous net reclassification improvement indices. **5d:** Compares the percentage of people reclassified to an appropriated KC risk group after adding the MTAG_PRS_ vs the UKB_PRS_ to a model with age, sex and 10 ancestral principal components.

## DISCUSSION

In this large multi-trait GWAS analysis we show that cutaneous melanoma, “all-cancer”, pigmentation traits, autoimmune diseases and other serum metabolic biomarkers are genetically correlated with BCC and SCC. We have leveraged this genetic correlation using the MTAG approach to identify 78 and 69 independent genome-wide significant loci for BCC and SCC risk respectively, the most common skin cancers among fair skinned people. 19 BCC and 15 SCC novel loci were replicated in the 23andMe cohort, indicating our study uncovers a number of novel findings relevant to keratinocyte cancer biology.

Firstly, we identify novel loci in the pigmentation pathways for both BCC and SCC susceptibility. Due to the importance of sun exposure in keratinocyte cancer biology ^106^, several new loci for BCC and SCC were linked to pigmentation traits including skin colour, red hair, skin tanning response and sunburns. The gene set analysis results also confirmed we identified biological pathways involved in melanin biosynthesis and DNA damage response.

Second, our study affirms the role of immune-regulatory processes and pathways in the BCC and SCC susceptibility. We show novel BCC and SCC loci known for immune regulatory processes including; HIV viral load modulation ^67,68^, innate immune response for coronaviruses (through *IFIH1*) ^107–109^, autoimmune disease susceptibility and medication use especially for thyroid or hypothyroidism. It indicates possible shared biology between keratinocyte cancers and immune-related viruses.

Third, immunosuppressive medication including azathioprine and cyclosporin A have been implicated in BCC and SCC risk ^110,111^. While we uncovered novel KC loci linked to immune-related medication use including; anti-asthmatic inhalants, and thyroid preparations ^63^, it is likely that medication-related loci underpinned here are just a proxy indicator for the autoimmune disease. Thus, these medications are unlikely to cause BCC or SCC. In addition, even if these diseases were all treated with drugs that greatly increased the risk of KC they are (a) too rare to lead to a cryptic genetic correlation as large as what we see here e.g. for hypothyroidism (*rg* = -0.19, *P* = 1.05 × 10^-4^) (**Supplementary Table 1**), and (b) the genetic correlation e.g. for hypothyroidism was negative with BCC where a drug induced cryptic overlap would give a positive genetic correlation.

Fourth, our study also highlights the potential role of cardiometabolic biomarkers in BCC/SCC risk. Besides the PUFA levels, whose causal association link with the BCC risk has been established through a Mendelian randomisation study ^112^, our results highlight a potential causal relationship between cardiometabolic biomarkers including; diastolic and systolic blood pressure, lipids, serum glucose, cholesterol and adiposity, and the risk of BCC and SCC. As it is the case for PUFA, downstream metabolism of these cardiometabolic biomarkers such as lipids and cholesterol results in oncogenic inflammatory biomarkers (e.g. prostaglandins E, thromboxane A2, and leukotriene B). However, some novel variants for the cardiometabolic pathway could be influencing BCC and SCC risk through already known pigmentation and immune regulatory biological pathways e.g. rs1136165 in *CKB*, and rs10774625 in *ATXN2* ^31,32,76,97^.

Fifth, we also unveil important genes with a potential role in BCC and SCC initiation and progression e.g. *FAP, CDKL1, MARK3, RAB11FIP2, GAB2, SUOX* and *SOX6*. Although loci within these genes had pleiotropic effects with pigmentation traits, the aforementioned genes have established roles in tumor cell proliferation, migration and invasion, and downregulation of apoptosis in a number of cancers e.g. melanoma, colorectal cancer, and breast cancer ^39, 48–50,74,75,113^. Besides the novel findings, our results further emphasise the shared biology between cutaneous melanoma and keratinocyte cancers. In total four novel loci for BCC and SCC at *ATM, DSTYK, GPR98,* and *SOX6* were previously known for CM ^20,103,104^.

One strength of the MTAG method is the increase in statistical power to identify several loci that a standard single trait GWAS would not have done. For example, using MTAG we increased our sample size by 2.6 times and 8.3 times for BCC and SCC respectively. Owing to the great improvement in statistical power, our MTAG-derived BCC PRS outperformed (for KC risk stratification) the one derived from a single trait BCC GWAS.

One caveat with the MTAG approach is that it assumes that the genetic variants have a homogeneous effect across all the included traits so that the results are not driven by a certain trait to result in false positives ^15^. Firstly, when we compared the genetic correlation and the MTAG results before and after excluding genomic regions with very large effect sizes including; *HLA, ASIP, IRF4, MC1R, SLC45A2*, and *CDKN2A,* the MTAG results were essentially unchanged (BCC; 78 vs 66 loci; SCC: 69 vs 64 loci; and **Supplementary Figure 1**) and there was a high concordance (**Supplementary Figure 2**). Secondly, there was good replication of our results in an independent cohort, which counters concerns of false positives.

In conclusion, leveraging the genetic correlation between skin cancers, autoimmune diseases and pigmentation traits revealed novel susceptibility loci for SCC and BCC, as well as produced a powerful and optimised PRS for KC risk stratification. Novel loci are implicated in keratinocyte cancer development and progression, pigmentation, cardiometabolic pathways, and immune-regulatory pathways including; innate immunity against coronaviruses, HIV-1 viral load modulation and disease progression.

## METHODS AND MATERIALS

### Cohorts

#### Discovery cohorts

Participants that contributed to the phenotype-specific genome-wide association studies were of homogenous European ancestry drawn from different cohorts from Australia, Europe and America. While there was sample overlap across the included GWAS, MTAG adjusts and corrects for biases due to sample overlap ^15^. The major cohorts used included; the UK Biobank (UKB) ^18,19^, QSkin Sun and Health Study (QSkin) (Olsen et al. 2012), eMERGE (dbGaP, study accession: phs000360.v3.p1) and GERA (dbGaP, study accession: phs000674.v3.p3), a melanoma meta-analysis consortium (**Supplementary Table 9**) ^20^, as well as publicly available GWAS summary statistics from international cohorts and consortium. Details for each cohort including ethics oversight are described in the **Supplementary Information**.

#### Replication cohort: 23andMe Research Cohort

23andMe, Inc. is a direct-to-consumer genetic company that collected both self-reported phenotypes and genetic data from participants who provided informed consent and participated in the research online, under a protocol approved by the external Association for the Accreditation of Human Research Protection Program (AAHRPP)-accredited Institutional Review Board (IRB), Ethical & Independent Review Services (E&I Review). The BCC cohort included 2,523,630 participants of European ancestry; 251,963 BCC cases and 2,271,667 controls, and 44.65% males. The SCC dataset included 2,529,399 participants of European ancestry; 134,700 SCC cases and 2,394,699 controls, and 44.65% males. Further details on data collection, validation, genotyping, imputation and quality control have been published before ^8,114^.

### BCC PRS application cohort: The Canadian Longitudinal Study on Aging (CLSA)

The Canadian Longitudinal Study on Aging (CLSA) is a prospective large population-based cohort in Canada comprising about 50,000 participants (45-85 years) randomly recruited between 2010 and 2015 from ten provinces ^115,116^. More information about the cohort has been published elsewhere ^115,116^ and summarised here. It consists of two cohorts; the “Tracking cohort” of ∼ 20,000 participants recruited through a telephone questionnaire in ten provinces, and the “Comprehensive cohort” with ∼ 30,000 individuals who provided data through an in-person questionnaire, clinical/physical tests and biological samples (e.g for genetic data) in seven provinces.

In general, at baseline information on relevant variants including age, and sex were recorded, and participants were also asked whether they had been diagnosed with any cancer including KC (yes/no) by a health professional. Between 2015 and 2018 the first follow up assessment was conducted and participants were asked again if they had been diagnosed with cancer, and KC during the follow up period. Thus, the CLSA dataset we used included; the “Baseline Comprehensive Dataset version 4.0” and “Follow-up 1 Comprehensive Dataset version 1.0”. At the time of analysis, ∼ 30,000 individuals had genetic data available, genotyped using 820K UK Biobank Axiom Array (Affymetrix)^115^, and imputed using the TopMed imputation server^117^. The CLSA is overseen by the Canadian Institutes of Health Research (CIHR) and its protocol has been reviewed and approved by 13 research ethics boards in Canada. All participants provided written informed consent.

Firstly, for purposes of validation and selection of the optimal PRS models (as described below in **Stage 6** analysis) we randomly selected 1,523 cancer-free controls and 388 prevalent KC cases at the baseline. Thus, our validation sample included 1,911 participants with a mean age of 65.81 years (sd =10.25) and 52.75% males.

Secondly, for we tested the BCC PRSs a second sample (unrelated to the validation dataset) of 18,933 participants of European ancestry, with a mean age of 61.80 years (sd = 9.84), followed up for a mean duration of 2.9 years (sd = 0.3) and 49.63% males. Only participants with complete data on age, sex, and cancer status and KC diagnosis were included. Thus, 18,139 controls with no history of any cancer (at follow up 1) and 794 participants who developed KC during follow up.

### Statistical analysis

#### Stage 1: GWAS for BCC, SCC and related traits

We conducted two case-control GWAS using UKB data for BCC, N = 307,684 (20,791 cases and 286,893 controls) and SCC, N = 294,294 (7,402 SCC cases and 286,892 controls) of European ancestry. We adjusted for age and sex as well as the first ten ancestral principal components (PCs) in order to control for biases from population stratification. We used Scalable and Accurate Implementation of GEneralized mixed model (SAIGE) software for the analysis since it controls for sample relatedness and case-control imbalance ^25^. Analysis was restricted to single nucleotide polymorphism (SNPs) with minor allele frequency (MAF) > 1% and an imputation quality score of 0.3. BCC/SCC. Case ascertainment and definition are described in **Supplementary Information.** In addition, we conducted GWAS for pigmentation traits (e.g. skin colour, hair colour, tanning response, skin burn, sunburn, etc.), all-cancer, autoimmune conditions, and blood biochemistry biomarkers (e.g. C-reactive protein, vitamin D, glucose, albumin, aspartate aminotransferase, gamma-glutamyl transferase, etc.) using data from international cohorts including; UKB, QSkin, and GERA as described in **Supplementary Information, Supplementary Table 2 and Supplementary Table 3.** We also conducted GWAS on KC and all-cancer after accessing data from eMERGE (dbGaP, study accession: phs000360.v3.p1) and GERA (dbGaP, study accession: phs000674.v3.p3) cohorts respectively **(Supplementary Information).** We also accessed publicly available GWAS summary statistics e.g. for cutaneous melanoma ^20^, smoking ^28^, education attainment ^27^, body mass index ^118^, hypothyroidism, type 1 diabetes, and rheumatoid arthritis ^25^, vitiligo ^26^ **(Supplementary Information, Supplementary Table 2 and Supplementary Table 3)**

#### Stage 2: Genetic correlation between BCC, SCC and related traits

We used LDSC version 1.0.1 ^119^, to compute the genetic correlation (*r_g_*) ^16^ between BCC and a range of other traits including; other skin cancer types, pigmentation traits, autoimmune traits and biochemistry biomarkers (recently released in the UKB). We then repeated this process for SCC instead of BCC. We used data from publicly available GWAS, as well as GWAS data from international cohorts of participants of European ancestry (conducted in stage 1 above). Traits with a statistically significant (*P* < 0.05) *r_g_* greater than 10% with either BCC or SCC were selected and included in the MTAG model (**Figure 1** **& Supplementary Table 1**). We further sought for additional traits that were genetically correlated with BCC or SCC using data from the LD hub catalog ^17^. Out of about 700 phenotypes, no additional phenotypes were selected to be included in the final MTAG model.

In total 22 traits including the initial input BCC and SCC GWAS from different cohorts of European ancestry met the inclusion criteria. The 22 genetically correlated traits included; BCC, SCC, skin colour, hair colour excluding red hair, hypothyroidism, type 1 diabetes, gamma-glutamyltransferase, aspartate aminotransferase, serum vitamin D levels, albumin, C-reactive protein, and glucose in the UK Biobank ^19^, KC, red hair and mole count in the QSkin ^21^, KC in eMERGE (dbGaP, study accession: phs000360.v3.p1), all-cancer in GERA cohort (dbGaP, study accession: phs000674.v3.p3), melanoma risk as measured by the latest and largest melanoma risk gwas meta-analysis ^20^, vitiligo ^26^, education attainment ^27^ and smoking ^28^. All the above studies excluded 23andMe, to enable us to utilise the 23andMe data as a replication set. Details on the phenotypic measurements and definitions are described in the **Supplementary Information, Supplementary Table 2 and Supplementary Table 3**

#### Stage 3: Multi-trait analysis of GWAS summary statistics

Next, using a total of 22 genetically correlated traits we conducted a multi-phenotype analysis of GWAS summary statistics (generated at stage 1 analysis and selected in stage 2) using MTAG software version 1.0.8 ^15^. MTAG default settings were used. MTAG combines GWAS summary statistics for genetically correlated traits into a meta-analysis while accounting for genetic correlation, sample overlap, maximising power to identify loci associated with the trait(s) of interest (here BCC and SCC) ^15^. MTAG generates trait specific results for each phenotype included in the model. BCC and SCC GWAS summary data from UKB from stage 1 were included as trait 1 and 2 respectively in the model below;

*MTAG model: BCC + SCC + melanoma + pigmentation traits + autoimmune traits + …….+ trait n.*

After the quality control measures, the analysis was restricted to 5,301,239 SNPs common in all the 22 GWAS with a minor allele frequency of >1%, and no ambiguous alleles. We assessed the effective increase in sample size after MTAG by comparing the average chi-squared before and after MTAG for BCC and for SCC using the following formula:

*(1-average_chi_MTAG_output)/(1-average_chi_MTAG_input)*

Where MTAG input corresponds to the input for either BCC or SCC in the UKB dataset.

We took forward the a) BCC and b) SCC MTAG output summary statistic results for further post-gwas analysis in stage 4 and replication in stage 5. BCC and SCC Manhattan plots, and Q-Q plots are presented in **Figures 2, and** **Figure 3, and in Supplementary Figure 1 and Supplementary Figure 2.**

#### Stage 3.1 Sensitivity analyses

MTAG assumes a homogeneous effect across all the included traits ^15^. However, due to their strong association with some input traits the following genomic regions were removed; *CDKN2A, SLC45A2*, *IRF4* and *HLA* for autoimmune, and *ASIP* and *MC1R* for pigmentation or CM, violate this assumption. We conducted sensitivity analyses excluding these regions before implementing our MTAG model. Using the stage 1 BCC GWAS summary statistics, we removed extended regions for *ASIP* on chromosome 20 (30-36 mega bases (mb)), *MHC* regions on chromosome 6 (25 - 36 mb), and *MC1R* on chromosome 16 (87-90.3 mb). We also removed 2mb around the most significant SNP in the following regions; rs12203592 (6:396321) in the *IRF4* region on chromosome 6, rs3731239 (9:21974218) in the *CDKN2A* region on chromosome 9, and rs16891982 (5:33951693) in the *SLC45A2* region on chromosome 5. We compared the genetic correlation between BCC/SCC before and after removing the genomic regions with known strong associations and high LD (**Supplementary Figure 2**), before running the full MTAG model of 22 traits described above. The MTAG results with and without the above genomic regions were also compared (**Supplementary Figure 1**).

#### Stage 4: Post-GWAS analysis

We used FUMA v.1.3.6 ^120^, to identify independent, genome wide significant SNPs and the genomic risk loci, and performed annotation of candidate SNPs in the genomic loci and functional gene mapping. We also conducted gene-based and pathway analyses using MAGMA v.1.7, as implemented in FUMA v.1.3.6 ^121^. For the gene pathway analysis, gene ontology (GO) and curated gene sets from MSigDB (v5.2) ^122^ were used and corrected for multiple testing. GWAS catalog ^123^ and Open Targets platform ^124^ were used to annotate novel loci and their relationship with other traits.

#### Stage 5: Replication of the BCC and SCC MTAG results

Next, we sought to replicate the BCC and SCC susceptibility loci in a large independent cohort using data from the 23andMe research cohort. For BCC the replication cohort included 251,963 self-reported cases and 2,271,667 controls while the SCC replication comprised 134,700 cases and self-reported cases and 2,394,699 controls of European ancestry filtered to remove close relatives.

Previous studies have shown high accuracy of 23andMe BCC/SCC self-reported cases ^8^ and high genetic correlation (*r_g_* > 0.9) between the histologically confirmed UKB BCC/SCC data and 23andMe data ^13^. Age, sex, and population stratification using five PCs were adjusted for in both analyses in a logistic regression i.e.

*BCC or SCC ∼ genotype + age + sex + pc.0 + pc.1 + pc.2 + pc.3 + pc.4 + v2_platform + v3_0_platform + v3_1_platform + v4_platform.*

The BCC results were adjusted for a genomic control inflation factor λ=1.286. The equivalent inflation factor for 1000 cases and 1000 controls λ1000=1.001, and for 10000, λ10000=1.006. In a similar way, the SCC results were adjusted for a genomic control inflation factor λ=1.172. The equivalent inflation factor for 1000 cases and 1000 controls λ1000=1.001, and for 10000, λ10000=1.007.

We compared the concordance of the effect sizes (log OR) for the MTAG results versus the replication results (**Figure 4c** and **Figure 4d**). We further analysed the number of loci that replicated at genome-wide significant level (*P* = 5.0 x 10^-8^), after multiple testing correction (i.e. Bonferroni correction *P* = 6.49 x 10^-4^ for BCC and *P* = 7.24 x 10^-4^ for SCC) and at a nominal *P* = 0.05.

#### Stage 6: Validation of the BCC Polygenic Risk Score in a selected sample of participants in CLSA

To construct two comparable polygenic risk scores (PRSs) for BCC, we separately used the BCC MTAG output (generated in stage 3) and the UKB BCC single-trait GWAS (generated in stage 1) summary statistics as the discovery data sets. MTAG ^15^ drops SNPs with extremely significant associations with any input trait, which resulted in a number of previously reported pigmentation associated SNPs being dropped from the model. Hence in both the MTAG and UKB discovery GWAS summary statistics we also included four functional SNPs (rs1805007 for *MC1R*, rs1126809 for *TYR*, rs6059655 for *ASIP*, and rs12203592 for *IRF4*) that would otherwise have been dropped in the PRS using the weights from a previously published BCC PRS ^125^.

Next using autosomal, non-ambiguous, and bi-allelic SNPs overlapping in the CLSA cohort (MTAG discovery = 5,300,872 SNPs and UKB discovery = 5,300,868 SNPs) we performed LD clumping based on (r^2^=0.005 and LD window= 5000 kb, *P* =1) to yield 62,494 and 62,884 independent SNPs for MTAG_PRS_ and UKB_PRS_ models respectively. PLINK 1.90b6.8 ^126^ for clumping. Using the clumped independent SNPs above, we generated PRS models at varying p-value thresholds i.e. S1 (P < 5×10^-8^), S2 (P < 10^-7^), S3 (P < 10^-6^), S4 (P < 10^-5^), S5 (P < 10^-4^), S6 (P < 10^-3^), S7 (P < 10^-2^) and S8 (P < 10^-1^) in validation sample of 1,911 participants split from the CLSA cohort using log odds ratio (from the respective discovery GWAS; MTAG or UKB) as weights. PLINK2 (v2.00a3LM 5 May 2021 release) ^126^ was used for generating the PRS scores. For both MTAG and UKB PRS models, we used Nagelkerke’s R^2 127^, a metric for model fitness used for selecting the optimal model. We computed the R^2^ by comparing the model fitness between models with PRSs (*BCC∼MTAG_PRS_ or UKB_PRS_ + age + sex + 10 Pcs*) and a null model using predictABEL package ^128^ in R software version 4.0.2 ^129^.

#### Stage 7: BCC Polygenic Risk Score and Keratinocyte Cancer Risk Prediction in the Canadian Longitudinal Study of Aging

To determine the ability of our MTAG GWAS data to predict skin cancer, we used 18,933 participants of European ancestry with data on KC risk in the Canadian Longitudinal Study of Aging (CLSA). We included 18,139 controls with no history of any cancer (both at baseline and follow-up) and 794 cases who developed KC during the 2.9 years (on average) of follow-up following baseline recruitment. Separate BCC and SCC data were unavailable in this cohort, and as 80% of KC cases are BCC cases ^130^ we tested the performance MTAG_PRS_ vs UKB_PRS_ derived for BCC.

Using PLINK2 (v2.00a3LM 5 May 2021 release) ^126^, we generated individual scores for CLSA participants for both the BCC MTAG_PRS_ and UKB_PRS_ weighted by their respective effect sizes (log odds ratios). The genetic scores were standardized to variance of 1 in order to interpret the associations as odds ratio per standard deviation increase in the PRS. We compared the performance of the two BCC PRSs (MTAG_PRS_ vs UKB_PRS_) based on the magnitude of the association (odds ratios) and the net reclassification improvement for KC risk using R version 4.0.2^129^. For net reclassification improvement, we compared the net reclassification index and the percentage of the participants who got reclassified to an appropriate risk group/tertile i.e. the low risk (bottom tertile), moderate risk (middle tertile), and high risk (top tertitle) after adding the MTAG_PRS_ vs UKB_PRS_ to the base model containing age, sex and the 10 PCs.

## DATA AVAILABILITY

### GWAS summary statistics

The full GWAS summary statistics for this study will be made available through the NHGRI-EBI GWAS Catalogue (https://www.ebi.ac.uk/gwas/downloads/summary-statistics).

### Polygenic risk scores

The PRS developed and utilised in this study are provided in the **Supplementary Tables 7 and 8** and will also be made available via the PRS catalog https://www.pgscatalog.org/

### UK Biobank

UK Biobank data is available through application via https://www.ukbiobank.ac.uk/. Information on UKB blood biochemistry biomarkers can be found in the UKB dataset: http://biobank.ctsu.ox.ac.uk/crystal/label.cgi?id=17518

### Canadian Longitudinal Study on Aging

Data are available from the Canadian Longitudinal Study on Aging (www.clsa-elcv.ca) for researchers who meet the criteria for access to de-identified CLSA data.

### 23andMe

The variant-level data for the 23andMe replication dataset are fully disclosed in the manuscript.

### dbGaP

https://www.ncbi.nlm.nih.gov/gap/advanced_search/

## ONLINE RESOURCES AND SOFTWARE

**FUMA:** https://fuma.ctglab.nl/

**LD hub catalog:** http://ldsc.broadinstitute.org/ldhub/

**LDpair Tool:** https://ldlink.nci.nih.gov/?tab=ldpair

**Open target genetics:** https://genetics.opentargets.org/

**GWAS catalog:** https://www.ebi.ac.uk/gwas/

## Data Availability

GWAS summary statistics
The full GWAS summary statistics for this study will be made available through the NHGRI-EBI
GWAS Catalogue upon publication (https://www.ebi.ac.uk/gwas/downloads/summary-statistics).
Polygenic risk scores
The PRS developed and utilised in this study are provided in the Supplementary Tables 7 and 8.
UK Biobank
UK Biobank data is available through application via https://www.ukbiobank.ac.uk/.
Information on UKB blood biochemistry biomarkers can be found in the UKB dataset:
http://biobank.ctsu.ox.ac.uk/crystal/label.cgi?id=17518
Canadian Longitudinal Study on Aging
Data are available from the Canadian Longitudinal Study on Aging (www.clsa-elcv.ca) for
researchers who meet the criteria for access to de-identified CLSA data.
23andMe
The variant-level data for the 23andMe replication dataset are fully disclosed in the manuscript.
dbGaP
https://www.ncbi.nlm.nih.gov/gap/advanced_search/

## ACKNOWLEDGMENTS

The study was supported by a program grant (APP1073898) and a project grant (APP1063061) from the Australian National Health and Medical Research Council (NHMRC). MS was supported by the Australian Government Research Training Program (RTP) and the Faculty of Health Scholarship at Queensland University of Technology, Australia. SM and DCW are supported by Research Fellowships from the NHMRC. This study was conducted using data from UK Biobank (application number 25331), QSkin Sun and Health Study (Australia), Canadian Longitudinal Study on Aging (CLSA), the 23andMe Research Team (USA), eMERGE (USA), GERA (USA) and other publicly available data from other consortia. We thank the research participants and the employees of 23andMe for making this work possible.

The opinions expressed in this manuscript are the author’s own and do not reflect the views of the Canadian Longitudinal Study on Aging or any affiliated institution. This research was made possible using the data/bio-specimens collected by the Canadian Longitudinal Study on Aging (CLSA). Funding for the Canadian Longitudinal Study on Aging (CLSA) is provided by the Government of Canada through the Canadian Institutes of Health Research (CIHR) under grant reference: LSA 94473 and the Canada Foundation for Innovation. This research has been conducted using the CLSA dataset [Baseline Comprehensive Dataset version 4.0, Follow-up 1 Comprehensive Dataset version 1.0], under Application Number 190225. The CLSA is led by Drs. Parminder Raina, Christina Wolfson and Susan Kirkland.

The following members of the 23andMe Research Team contributed to this study: Stella Aslibekyan, Adam Auton, Elizabeth Babalola, Robert K. Bell, Jessica Bielenberg, Katarzyna Bryc, Emily Bullis, Daniella Coker, Gabriel Cuellar Partida, Devika Dhamija, Sayantan Das, Sarah L. Elson, Teresa Filshtein, Kipper Fletez-Brant, Pierre Fontanillas, Will Freyman, Pooja M. Gandhi, Karl Heilbron, Barry Hicks, David A. Hinds, Ethan M. Jewett, Yunxuan Jiang, Katelyn Kukar, Keng-Han Lin, Maya Lowe, Jey McCreight, Matthew H. McIntyre, Steven J. Micheletti, Meghan E. Moreno, Joanna L. Mountain, Priyanka Nandakumar, Elizabeth S. Noblin, Jared O’Connell, Aaron A. Petrakovitz, G. David Poznik, Morgan Schumacher, Anjali J. Shastri, Janie F. Shelton, Jingchunzi Shi, Suyash Shringarpure, Vinh Tran, Joyce Y. Tung, Xin Wang, Wei Wang, Catherine H. Weldon, Peter Wilton, Alejandro Hernandez, Corinna Wong, Christophe Toukam Tchakouté

## AUTHOR CONTRIBUTIONS

Conceptualization: MS, MHL, SM; Data curation: MS, MHL, DCW, CMO, SM; Formal analysis: MS; Funding acquisition: SM, MHL, DCW; Investigation: MS, MHL, DCW, CMO,SM; Methodology: MS, MHL, SM; Project administration: MS, MHL, SM; Resources: MHL, DCW, SM; Software: MS; Supervision: SM, MHL; Visualization: MS; Writing - original draft preparation: MS; Writing - revision of subsequent drafts: MS, SM, MHL, JSO, PG, DCW, CMO, PF. All authors contributed to the final version of the manuscript.

## CONFLICT OF INTEREST

Pierre Fontanillas, Chao Tian, and the 23andMe Research Team are employed by and hold stock or stock options in 23andMe, Inc. The rest of the authors declare no conflict of interests.

## Notes

### Author Declarations

The Human Research Ethics Committee of QIMR Berghofer Medical Research Institute gave ethical approval for this work.

## REFERENCES

1. Guy, G. P., Jr, Machlin, S. R., Ekwueme, D. U. & Yabroff, K. R. Prevalence and costs of skin cancer treatment in the U.S., 2002-2006 and 2007-2011. Am. J. Prev. Med. 48, 183–187 (2015).

2. Australian Institute of Health and Welfare. Cancer in Australia: Actual incidence data from 1982 to 2013 and mortality data from 1982 to 2014 with projections to 2017. Asia Pac. J. Clin. Oncol. 14, 5–15 (2018).

3. Fransen, M. et al. Non-melanoma skin cancer in Australia. Med. J. Aust. 197, 565–568 (2012).

4. Nagarajan, P. et al. Keratinocyte Carcinomas: Current Concepts and Future Research Priorities. Clin. Cancer Res. 25, 2379–2391 (2019).

5. Pandeya, N., Olsen, C. M. & Whiteman, D. C. The incidence and multiplicity rates of keratinocyte cancers in Australia. Med. J. Aust. 207, 339–343 (2017).

6. Zhu, K.-J. et al. Psoriasis regression analysis of MHC loci identifies shared genetic variants with vitiligo. PLoS One 6, e23089 (2011).

7. Coenen, M. J. H. et al. Common and different genetic background for rheumatoid arthritis and coeliac disease. Hum. Mol. Genet. 18, 4195–4203 (2009).

8. Chahal, H. S. et al. Genome-wide association study identifies 14 novel risk alleles associated with basal cell carcinoma. Nat. Commun. 7, 12510 (2016).

9. Hinks, A. et al. Investigation of type 1 diabetes and coeliac disease susceptibility loci for association with juvenile idiopathic arthritis. Ann. Rheum. Dis. 69, 2169–2172 (2010).

10. Roberts, M. R., Asgari, M. M. & Toland, A. E. Genome-wide association studies and polygenic risk scores for skin cancer: clinically useful yet? Br. J. Dermatol. 181, 1146–1155 (2019).

11. Al Olama, A. A. et al. A meta-analysis of 87,040 individuals identifies 23 new susceptibility loci for prostate cancer. Nat. Genet. 46, 1103–1109 (2014).

12. Qian, X. et al. The Tensin-3 protein, including its SH2 domain, is phosphorylated by Src and contributes to tumorigenesis and metastasis. Cancer Cell 16, 246–258 (2009).

13. Liyanage, U. E. et al. Combined analysis of keratinocyte cancers identifies novel genome-wide loci. Hum. Mol. Genet. 28, 3148–3160 (2019).

14. Manousaki, D. et al. Genome-wide Association Study for Vitamin D Levels Reveals 69 Independent Loci. Am. J. Hum. Genet. 106, 327–337 (2020).

15. Turley, P. et al. Multi-trait analysis of genome-wide association summary statistics using MTAG. Nat. Genet. 50, 229–237 (2018).

16. Ni, G., Moser, G., Schizophrenia Working Group of the Psychiatric Genomics Consortium, Wray, N. R. & Lee, S. H. Estimation of Genetic Correlation via Linkage Disequilibrium Score Regression and Genomic Restricted Maximum Likelihood. Am. J. Hum. Genet. 102, 1185–1194 (2018).

17. Zheng, J. et al. LD Hub: a centralized database and web interface to perform LD score regression that maximizes the potential of summary level GWAS data for SNP heritability and genetic correlation analysis. Bioinformatics 33, 272–279 (2017).

18. Sudlow, C. et al. UK biobank: an open access resource for identifying the causes of a wide range of complex diseases of middle and old age. PLoS Med. 12, e1001779 (2015).

19. Bycroft, C. et al. The UK Biobank resource with deep phenotyping and genomic data. Nature 562, 203– 209 (2018).

20. Landi, M. T. et al. Genome-wide association meta-analyses combining multiple risk phenotypes provide insights into the genetic architecture of cutaneous melanoma susceptibility. Nat. Genet. 52, 494–504 (2020).

21. Olsen, C. M. et al. Cohort profile: the QSkin Sun and Health Study. Int. J. Epidemiol. 41, 929–929i (2012).

22. McCarty, C. A. et al. The eMERGE Network: a consortium of biorepositories linked to electronic medical records data for conducting genomic studies. BMC Med. Genomics 4, 13 (2011).

23. Kho, A. N. et al. Electronic medical records for genetic research: results of the eMERGE consortium. Sci. Transl. Med. 3, 79re1 (2011).

24. Banda, Y. et al. Characterizing Race/Ethnicity and Genetic Ancestry for 100,000 Subjects in the Genetic Epidemiology Research on Adult Health and Aging (GERA) Cohort. Genetics 200, 1285–1295 (2015).

25. Zhou, W. et al. Efficiently controlling for case-control imbalance and sample relatedness in large-scale genetic association studies. Nat. Genet. 50, 1335–1341 (2018).

26. Jin, Y. et al. Genome-wide association studies of autoimmune vitiligo identify 23 new risk loci and highlight key pathways and regulatory variants. Nat. Genet. 48, 1418–1424 (2016).

27. Okbay, A. et al. Genome-wide association study identifies 74 loci associated with educational attainment. Nature 533, 539–542 (2016).

28. Liu, M. et al. Association studies of up to 1.2 million individuals yield new insights into the genetic etiology of tobacco and alcohol use. Nat. Genet. 51, 237–244 (2019).

29. Owusu, M. et al. Mapping the Human Kinome in Response to DNA Damage. Cell Rep. 26, 555–563.e6 (2019).

30. Kato, T. et al. Isolation of a novel human gene, MARKL1, homologous to MARK3 and its involvement in hepatocellular carcinogenesis. Neoplasia 3, 4–9 (2001).

31. Kichaev, G. et al. Leveraging Polygenic Functional Enrichment to Improve GWAS Power. Am. J. Hum. Genet. 104, 65–75 (2019).

32. Morgan, M. D. et al. Genome-wide study of hair colour in UK Biobank explains most of the SNP heritability. Nat. Commun. 9, 5271 (2018).

33. Dong, W., Qin, G. & Shen, R. Rab11-FIP2 promotes the metastasis of gastric cancer cells. International journal of cancer 138, (2016).

34. Dong, W. & Wu, X. Overexpression of Rab11-FIP2 in colorectal cancer cells promotes tumor migration and angiogenesis through increasing secretion of PAI-1. Cancer Cell Int. 18, 35 (2018).

35. Visconti, A. et al. Genome-wide association study in 176,678 Europeans reveals genetic loci for tanning response to sun exposure. Nat. Commun. 9, 1684 (2018).

36. Adhikari, K. et al. A GWAS in Latin Americans highlights the convergent evolution of lighter skin pigmentation in Eurasia. Nat. Commun. 10, 358 (2019).

37. Endo, C. et al. Genome-wide association study in Japanese females identifies fifteen novel skin-related trait associations. Sci. Rep. 8, 8974 (2018).

38. Hysi, P. G. et al. Genome-wide association meta-analysis of individuals of European ancestry identifies new loci explaining a substantial fraction of hair color variation and heritability. Nat. Genet. 50, 652– 656 (2018).

39. Kondreddy, V., Magisetty, J., Keshava, S., Vijaya Mohan Rao, L. & Pendurthi, U. R. Gab2 (Grb2-Associated Binder2) Plays a Crucial Role in Inflammatory Signaling and Endothelial Dysfunction. Arterioscler. Thromb. Vasc. Biol. 41, 1987 (2021).

40. Ding, C.-B., Yu, W.-N., Feng, J.-H. & Luo, J.-M. Structure and function of Gab2 and its role in cancer (Review). Mol. Med. Rep. 12, 4007 (2015).

41. Bentires-Alj, M. et al. A role for the scaffolding adapter GAB2 in breast cancer. Nat. Med. 12, 114–121 (2006).

42. Brummer, T. et al. Increased proliferation and altered growth factor dependence of human mammary epithelial cells overexpressing the Gab2 docking protein. J. Biol. Chem. 281, 626–637 (2006).

43. Qin, Y.-R. et al. Characterization of tumor-suppressive function of SOX6 in human esophageal squamous cell carcinoma. Clin. Cancer Res. 17, 46–55 (2011).

44. Jiang, W. et al. Identification of Sox6 as a regulator of pancreatic cancer development. J. Cell. Mol. Med. 22, 1864–1872 (2018).

45. Li, Y., Xiao, M. & Guo, F. The role of Sox6 and Netrin-1 in ovarian cancer cell growth, invasiveness, and angiogenesis. Tumour Biol. 39, 1010428317705508 (2017).

46. Li, Y.-C. et al. MicroRNA-766 targeting regulation of SOX6 expression promoted cell proliferation of human colorectal cancer. Onco. Targets. Ther. 8, 2981–2988 (2015).

47. Guo, X., Yang, M., Gu, H., Zhao, J. & Zou, L. Decreased expression of SOX6 confers a poor prognosis in hepatocellular carcinoma. Cancer Epidemiol. 37, 732–736 (2013).

48. Song, Z., Lin, J., Sun, Z., Ni, J. & Sha, Y. RNAi-mediated downregulation of CDKL1 inhibits growth and colony-formation ability, promotes apoptosis of human melanoma cells. J. Dermatol. Sci. 79, (2015).

49. Qin, C. et al. CDKL1 promotes tumor proliferation and invasion in colorectal cancer. Onco. Targets. Ther. 10, 1613–1624 (2017).

50. Tang, L., Gao, Y., Yan, F. & Tang, J. Evaluation of cyclin-dependent kinase-like 1 expression in breast cancer tissues and its regulation in cancer cell growth. Cancer Biother. Radiopharm. 27, 392–398 (2012).

51. Hu, J. et al. FERM domain-containing protein FRMD5 regulates cell motility via binding to integrin β5 subunit and ROCK1. FEBS Letters vol. 588 4348–4356 (2014).

52. Wang, T. et al. FERM-containing protein FRMD5 is a p120-catenin interacting protein that regulates tumor progression. FEBS Lett. 586, 3044–3050 (2012).

53. Hoffmann, T. J. et al. Genome-wide association analyses using electronic health records identify new loci influencing blood pressure variation. Nat. Genet. 49, 54–64 (2017).

54. Nakamura, K. et al. SUOX is negatively associated with multistep carcinogenesis and proliferation in oral squamous cell carcinoma. Med. Mol. Morphol. 51, 102–110 (2018).

55. Jin, G.-Z. et al. SUOX is a promising diagnostic and prognostic biomarker for hepatocellular carcinoma. J. Hepatol. 59, 510–517 (2013).

56. Yano, Y. et al. Sulfite Oxidase Is a Novel Prognostic Biomarker of Advanced Gastric Cancer. In Vivo 35, 229–237 (2021).

57. Cui, D. et al. The ribosomal protein S26 regulates p53 activity in response to DNA damage. Oncogene 33, 2225–2235 (2014).

58. Sharma, M. D. et al. An inherently bifunctional subset of Foxp3+ T helper cells is controlled by the transcription factor eos. Immunity 38, 998–1012 (2013).

59. Pan, F. et al. Eos mediates Foxp3-dependent gene silencing in CD4+ regulatory T cells. Science 325, 1142–1146 (2009).

60. Onengut-Gumuscu, S. et al. Fine mapping of type 1 diabetes susceptibility loci and evidence for colocalization of causal variants with lymphoid gene enhancers. Nat. Genet. 47, 381–386 (2015).

61. Ferreira, M. A. et al. Shared genetic origin of asthma, hay fever and eczema elucidates allergic disease biology. Nat. Genet. 49, 1752–1757 (2017).

62. Laufer, V. A. et al. Genetic influences on susceptibility to rheumatoid arthritis in African-Americans. Hum. Mol. Genet. 28, 858–874 (2019).

63. Wu, Y. et al. Genome-wide association study of medication-use and associated disease in the UK Biobank. Nat. Commun. 10, 1891 (2019).

64. McLaren, P. J. et al. Polymorphisms of large effect explain the majority of the host genetic contribution to variation of HIV-1 virus load. Proc. Natl. Acad. Sci. U. S. A. 112, 14658–14663 (2015).

65. Ekenberg, C. et al. Association Between Single-Nucleotide Polymorphisms in HLA Alleles and Human Immunodeficiency Virus Type 1 Viral Load in Demographically Diverse, Antiretroviral Therapy-Naive Participants From the Strategic Timing of AntiRetroviral Treatment Trial. J. Infect. Dis. 220, 1325– 1334 (2019).

66. Hüttenrauch, F., Pollok-Kopp, B. & Oppermann, M. G protein-coupled receptor kinases promote phosphorylation and beta-arrestin-mediated internalization of CCR5 homo- and hetero-oligomers. J. Biol. Chem. 280, 37503–37515 (2005).

67. Alvarez, V., López-Larrea, C. & Coto, E. Mutational analysis of the CCR5 and CXCR4 genes (HIV-1 co-receptors) in resistance to HIV-1 infection and AIDS development among intravenous drug users. Hum. Genet. 102, 483–486 (1998).

68. Carrington, M., Dean, M., Martin, M. P. & O’Brien, S. J. Genetics of HIV-1 infection: chemokine receptor CCR5 polymorphism and its consequences. Hum. Mol. Genet. 8, 1939–1945 (1999).

69. Hartmann, T. N. et al. Human B cells express the orphan chemokine receptor CRAM-A/B in a maturation-stage-dependent and CCL5-modulated manner. Immunology vol. 125 252–262 (2008).

70. Kato, H. et al. Differential roles of MDA5 and RIG-I helicases in the recognition of RNA viruses. Nature 441, 101–105 (2006).

71. Wawrusiewicz-Kurylonek, N. et al. The interferon-induced helicase C domain-containing protein 1 gene variant (rs1990760) as an autoimmune-based pathology susceptibility factor. Immunobiology 225, 151864 (2020).

72. Dias Junior, A. G., Sampaio, N. G. & Rehwinkel, J. A Balancing Act: MDA5 in Antiviral Immunity and Autoinflammation. Trends Microbiol. 27, 75–85 (2019).

73. Puré, E. & Blomberg, R. Pro-tumorigenic roles of fibroblast activation protein in cancer: back to the basics. Oncogene 37, 4343–4357 (2018).

74. Chen, L., Qiu, X., Wang, X. & He, J. FAP positive fibroblasts induce immune checkpoint blockade resistance in colorectal cancer via promoting immunosuppression. Biochem. Biophys. Res. Commun. 487, 8–14 (2017).

75. Wen, X. et al. Fibroblast Activation Protein-α-Positive Fibroblasts Promote Gastric Cancer Progression and Resistance to Immune Checkpoint Blockade. Oncology Research Featuring Preclinical and Clinical Cancer Therapeutics vol. 25 629–640 (2017).

76. Pickrell, J. K. et al. Detection and interpretation of shared genetic influences on 42 human traits. Nat. Genet. 48, 709–717 (2016).

77. Devallière, J. & Charreau, B. The adaptor Lnk (SH2B3): an emerging regulator in vascular cells and a link between immune and inflammatory signaling. Biochem. Pharmacol. 82, 1391–1402 (2011).

78. Katayama, H. et al. Lnk prevents inflammatory CD8^+^ T-cell proliferation and contributes to intestinal homeostasis. Eur. J. Immunol. 44, 1622–1632 (2014).

79. Pereira, B. I. et al. Sestrins induce natural killer function in senescent-like CD8 T cells. Nat. Immunol. 21, 684–694 (2020).

80. Lanna, A. et al. A sestrin-dependent Erk-Jnk-p38 MAPK activation complex inhibits immunity during aging. Nat. Immunol. 18, 354–363 (2017).

81. Okada, Y. et al. Genetics of rheumatoid arthritis contributes to biology and drug discovery. Nature 506, 376–381 (2014).

82. Jin, Y. et al. Genome-wide association analyses identify 13 new susceptibility loci for generalized vitiligo. Nat. Genet. 44, 676–680 (2012).

83. Ward, L. D. & Kellis, M. HaploReg: a resource for exploring chromatin states, conservation, and regulatory motif alterations within sets of genetically linked variants. Nucleic Acids Res. 40, D930–4 (2012).

84. Goudy, K. et al. Human IL2RA null mutation mediates immunodeficiency with lymphoproliferation and autoimmunity. Clin. Immunol. 146, 248–261 (2013).

85. Liu, J. Z. et al. Association analyses identify 38 susceptibility loci for inflammatory bowel disease and highlight shared genetic risk across populations. Nat. Genet. 47, 979–986 (2015).

86. Tsoi, L. C. et al. Large scale meta-analysis characterizes genetic architecture for common psoriasis associated variants. Nat. Commun. 8, 15382 (2017).

87. Dillon, S. R. et al. Interleukin 31, a cytokine produced by activated T cells, induces dermatitis in mice. Nat. Immunol. 5, 752–760 (2004).

88. McKay, J. D. et al. Large-scale association analysis identifies new lung cancer susceptibility loci and heterogeneity in genetic susceptibility across histological subtypes. Nat. Genet. 49, 1126–1132 (2017).

89. Lemaitre, R. N. et al. Genetic loci associated with plasma phospholipid n-3 fatty acids: a meta-analysis of genome-wide association studies from the CHARGE Consortium. PLoS Genet. 7, e1002193 (2011).

90. Guan, W. et al. Genome-wide association study of plasma N6 polyunsaturated fatty acids within the cohorts for heart and aging research in genomic epidemiology consortium. Circ. Cardiovasc. Genet. 7, 321–331 (2014).

91. Azrad, M., Turgeon, C. & Demark-Wahnefried, W. Current evidence linking polyunsaturated Fatty acids with cancer risk and progression. Front. Oncol. 3, 224 (2013).

92. Klarin, D. et al. Genetics of blood lipids among ∼300,000 multi-ethnic participants of the Million Veteran Program. Nat. Genet. 50, 1514–1523 (2018).

93. Hoffmann, T. J. et al. A large electronic-health-record-based genome-wide study of serum lipids. Nat. Genet. 50, 401–413 (2018).

94. Warner, J. P., Leek, J. P., Intody, S., Markham, A. F. & Bonthron, D. T. Human glucokinase regulatory protein (GCKR): cDNA and genomic cloning, complete primary structure, and chromosomal localization. Mamm. Genome 6, (1995).

95. van der Harst, P. & Verweij, N. Identification of 64 Novel Genetic Loci Provides an Expanded View on the Genetic Architecture of Coronary Artery Disease. Circ. Res. 122, 433–443 (2018).

96. Sung, Y. J. et al. A Large-Scale Multi-ancestry Genome-wide Study Accounting for Smoking Behavior Identifies Multiple Significant Loci for Blood Pressure. Am. J. Hum. Genet. 102, 375–400 (2018).

97. Plagnol, V. et al. Genome-wide association analysis of autoantibody positivity in type 1 diabetes cases. PLoS Genet. 7, e1002216 (2011).

98. Sulem, P. et al. Two newly identified genetic determinants of pigmentation in Europeans. Nat. Genet. 40, 835–837 (2008).

99. Kerns, S. L. et al. Radiogenomics Consortium Genome-Wide Association Study Meta-Analysis of Late Toxicity After Prostate Cancer Radiotherapy. J. Natl. Cancer Inst. 112, 179–190 (2020).

100. Wu, X. et al. ATM phosphorylation of Nijmegen breakage syndrome protein is required in a DNA damage response. Nature vol. 405 477–482 (2000).

101. Bakkenist, C. J. & Kastan, M. B. DNA damage activates ATM through intermolecular autophosphorylation and dimer dissociation. Nature 421, 499–506 (2003).

102. Duffy, D. L. et al. Novel pleiotropic risk loci for melanoma and nevus density implicate multiple biological pathways. Nat. Commun. 9, 4774 (2018).

103. Barrett, J. H. et al. Genome-wide association study identifies three new melanoma susceptibility loci. Nat. Genet. 43, 1108–1113 (2011).

104. Ransohoff, K. J. et al. Two-stage genome-wide association study identifies a novel susceptibility locus associated with melanoma. Oncotarget 8, 17586–17592 (2017).

105. Schumacher, F. R. et al. Association analyses of more than 140,000 men identify 63 new prostate cancer susceptibility loci. Nat. Genet. 50, 928–936 (2018).

106. D’Orazio, J., Jarrett, S., Amaro-Ortiz, A. & Scott, T. UV Radiation and the Skin. Int. J. Mol. Sci. 14, 12222–12248 (2013).

107. Zheng, Y. et al. Severe acute respiratory syndrome coronavirus 2 (SARS-CoV-2) membrane (M) protein inhibits type I and III interferon production by targeting RIG-I/MDA-5 signaling. Signal Transduct Target Ther 5, 299 (2020).

108. Yin, X. et al. MDA5 Governs the Innate Immune Response to SARS-CoV-2 in Lung Epithelial Cells. Cell Rep. 34, 108628 (2021).

109. Maiti, A. K. The African-American population with a low allele frequency of SNP rs1990760 (T allele) in IFIH1 predicts less IFN-beta expression and potential vulnerability to COVID-19 infection. Immunogenetics 72, 387–391 (2020).

110. Jiyad, Z., Olsen, C. M., Burke, M. T., Isbel, N. M. & Green, A. C. Azathioprine and Risk of Skin Cancer in Organ Transplant Recipients: Systematic Review and Meta-Analysis. Am. J. Transplant 16, (2016).

111. Kuschal, C. et al. Skin cancer in organ transplant recipients: effects of immunosuppressive medications on DNA repair. Exp. Dermatol. 21, 2–6 (2012).

112. Seviiri, M. et al. Polyunsaturated fatty acid levels and the risk of keratinocyte cancer: A Mendelian Randomisation analysis. Cancer Epidemiology Biomarkers & Prevention cebp.1765.2020 (2021) doi:10.1158/1055-9965.epi-20-1765.

113. Dong, W., Li, H. & Wu, X. Rab11-FIP2 suppressed tumor growth via regulation of PGK1 ubiquitination in non-small cell lung cancer. Biochem. Biophys. Res. Commun. 508, 60–65 (2019).

114. Chahal, H. S. et al. Genome-wide association study identifies novel susceptibility loci for cutaneous squamous cell carcinoma. Nat. Commun. 7, 12048 (2016).

115. Raina, P. et al. Cohort Profile: The Canadian Longitudinal Study on Aging (CLSA). Int. J. Epidemiol. 48, 1752–1753j (2019).

116. Raina, P. S. et al. The Canadian longitudinal study on aging (CLSA). Can. J. Aging 28, 221–229 (2009).

117. Taliun, D. et al. Sequencing of 53,831 diverse genomes from the NHLBI TOPMed Program. Nature 590, 290–299 (2021).

118. Yengo, L. et al. Meta-analysis of genome-wide association studies for height and body mass index in ∼700000 individuals of European ancestry. Hum. Mol. Genet. 27, 3641–3649 (2018).

119. Bulik-Sullivan, B. et al. An atlas of genetic correlations across human diseases and traits. Nat. Genet. 47, 1236–1241 (2015).

120. Watanabe, K., Taskesen, E., van Bochoven, A. & Posthuma, D. Functional mapping and annotation of genetic associations with FUMA. Nat. Commun. 8, 1826 (2017).

121. de Leeuw, C. A., Mooij, J. M., Heskes, T. & Posthuma, D. MAGMA: generalized gene-set analysis of GWAS data. PLoS Comput. Biol. 11, e1004219 (2015).

122. Liberzon, A. et al. The Molecular Signatures Database Hallmark Gene Set Collection. Cell Systems vol. 1 417–425 (2015).

123. MacArthur, J. et al. The new NHGRI-EBI Catalog of published genome-wide association studies (GWAS Catalog). Nucleic Acids Res. 45, D896–D901 (2017).

124. Ghoussaini, M. et al. Open Targets Genetics: systematic identification of trait-associated genes using large-scale genetics and functional genomics. Nucleic Acids Res. 49, D1311–D1320 (2021).

125. Seviiri, M. et al. Polygenic Risk Scores Allow Risk Stratification for Keratinocyte Cancer in Organ-Transplant Recipients. J. Invest. Dermatol. 141, 325–333.e6 (2021).

126. Chang, C. C. et al. Second-generation PLINK: rising to the challenge of larger and richer datasets. Gigascience 4, 7 (2015).

127. Nagelkerke, N. J. D. A note on a general definition of the coefficient of determination. Biometrika vol. 78 691–692 (1991).

128. Kundu, S., Aulchenko, Y. S., van Duijn, C. M. & Janssens, A. C. J. W. PredictABEL: an R package for the assessment of risk prediction models. Eur. J. Epidemiol. 26, 261–264 (2011).

129. R Core Team. R: A Language and Environment for Statistical Computing. (R Foundation for Statistical Computing, 2020).

130. Ciążyńska, M. et al. The incidence and clinical analysis of non-melanoma skin cancer. Sci. Rep. 11, 4337 (2021).

