## Supplementary Information for "A multi-phenotype analysis reveals 19 novel susceptibility loci for basal cell carcinoma and 15 for squamous cell carcinoma"

**Affiliations**

### ACRONYMS

Basal cell carcinoma (BCC), Genome wide association studies (GWAS), International Classification of Diseases (ICD), mega bases (mb), Keratinocyte cancers (KC), Linkage disequilibrium (LD), LD score regression (LDSC), Multi-trait analysis of GWAS (MTAG), Minor allele frequency (MAF), Polygenic risk score (PRS), Principal components (PCs), QSkin Sun and Health Study (QSkin), Scalable and Accurate Implementation of GEneralized mixed model (SAIGE), Squamous cell carcinoma (SCC), Standard deviation (SD), UK Biobank (UKB)

### METHODS

#### The UK Biobank and the respective GWAS

The UK Biobank is a population-based cohort comprising half a million adult participants (40-69 years) recruited between 2006 and 2010 from the United Kingdom. More information on participant recruitment, phenotype measurements, genotyping and participant follow up have been published elsewhere [1,2] and summarised here. The study was approved by the National North West Multi-Centre Research Ethics Committee (ref. 11/NW/0382) in the United Kingdom and all participants provided written informed consent. More information about ethics oversight in the UK Biobank can be accessed at https://www.ukbiobank.ac.uk/ethics/.

In the UKB, BCC and SCC cases were ascertained through linkage of participant hospital data with cancer registries in the UK. In this present study, we selected histo-clinically confirmed BCC and SCC cases based on International Classification of Diseases (ICD) 10 codes (UKB data field 40006) or ICD 9 codes (data field 40013) and histological confirmation (data field 40011) based on the ICD for Oncology, 3rd edition codes for SCC (8070, 8071, 8072, 8073, 8074, 8075, 8076, 8078) and BCC (8090, 8091, 8092, 8093, 8094, 8097, 8098). We excluded cancer *in-situ* and self-reported cases for SCC and BCC. Controls had no history of any cancer diagnosis. We conducted the case-control GWAS for BCC (20,791 cases and 286,893 controls) and SCC (7,402 SCC cases and 286,892 controls) as described following in the **Methods**.

In addition, we conducted GWAS on pigmentation related traits in the UKB using the following phenotypes; facial aging (data field 1757), skin colour (data field 1717), childhood sunburns (data field 1737) and hair colour excluding red hair (data field 1747). We fitted linear mixed models using BOLT-LMM v2.3 [3] adjusting for age, age$^2$, sex and the interaction between them, and 10 PCs. For each phenotype we arranged its categorical values/codes in the exact order described in **Supplementary Table 2** and **Supplementary Table 3** then we applied rank normalisation (for each phenotype) prior to running linear models. For example skin colour was ordered as; very fair, fair, light olive, dark olive, and brown while hair colour was; blonde, light brown, dark brown, and black. Red hair (data field 1747) was split off into a binary phenotype (red vs all others) and we conducted a case-control GWAS (14,354 cases, and 417,860 controls).

Using linear mixed modelling, we conducted quantitative GWAS on the following recently released blood biomarkers in the UKB; alanine aminotransferase (data field 30620), albumin (data field 30600), alkaline phosphatase (data field 30610), apolipoprotein A (data field 30630), apolipoprotein B (data field 30640), aspartate aminotransferase (data field 30650), C-reactive protein (data field 30710), calcium (data field 30680), cholesterol (data field 30690), creatinine (data field 30700), cystatin C (data field 30720), direct bilirubin (data field 30660), gamma glutamyltransferase (data field 30730), glucose (data field 30740), glycated haemoglobin (HbA1c) (data field 30750), High-density lipoprotein (HDL) cholesterol (data field 30760), insulin-like growth factor 1 (IGF-1) (data field 30770), low-density lipoprotein (LDL) direct (data field 30780), lipoprotein A (data field 30790), oestradiol (data field 30800), phosphate (data field 30810), sex hormone-binding globulin (SHBG) (data field 30830), testosterone (data field 30850), total bilirubin (data field 30840), total protein (data field 30860), triglycerides (data field 30870), urate (data field 30880), urea (data field 30670), and Vitamin D (data field 30890). We adjusted for age, sex and the first ten principal components. We included only participants of European ancestry and BOLT-LMM v2.3 [3] was used for the analysis. Sample sizes and details of each phenotype measurement are presented in **Supplementary Table 2**, and **Supplementary Table 3**. SNPs with MAF > 1% and imputation score greater than 0.3 were retained for subsequent analyses (**Methods**).

**The QSkin Sun and Health Study (QSkin) and the respective GWAS**

The QSkin Sun and Health Study (QSkin) has been extensively described elsewhere [4], and summarised here. Briefly, it is an Australian population based prospective cohort comprising over 43,000 adult participants aged 40-60 years recruited in 2011 with both clinical and skin-related trait data [4]. Participants residing in the state of Queensland were randomly sampled from the electoral rolls. In 2017 about 17,000 participants were genotyped using Illumina GSA arrays and imputed to the Haplotype Reference Consortium (HRC) panel version r1.1 2016 [5] using the Michigan Imputation Server [6]**.** Prior to imputation, genotype quality checks were conducted to remove individuals with high genotype missingness (>3%), relatedness (pihat >0.1875) and ancestry divergent from a European reference panel (> 6 SD on PC1 or 2 from European 1000 genome samples). In addition, SNPs with a MAF < 1% and Hardy-Weinberg equilibrium (HWE) p-value $< 1 \times 10^{-6}$ were excluded. After imputation, only SNPs with imputation quality score > 0.3 were retained. QSkin was approved by the Human Research Ethics Committee at QIMR Berghofer Medical Research Institute, Brisbane, Australia. All study participants provided written informed consent.

Data on skin and hair pigmentation traits, and skin cancers including KC were collected. KC (BCC and SCC) cases were ascertained through linkage of participant data with their health records in Medicare Australia and other Australian pathology registers [4]. Controls had no history of self-reported KC or actinic keratoses.

For ordinal traits we applied rank normalisation, and conducted GWAS using generalised linear models for mole count excluding melanoma cases (1=no moles, 2=some moles, 3=a few moles, and 4=many moles), skin burn type (1=not burn, 2=burn a little, 3=burn moderately, and 4=burn badly), skin colour (1=fair, 2=medium 3=olive/dark, and 4=black), skin tanning response (1=not tan, 2=tan lightly, 3=tan moderately, 4=tan deeply), hair colour excluding red (1=blonde, 2=light brown, 3=dark brown, and 4=black), and freckles (1=no freckles, 2=few freckles, 3=some freckles, and 4=many freckles). In addition, we conducted case-control GWAS for KC (8145 cases, 4797 controls), and red hair (973 cases, 15,202 controls). In all the GWAS above, we adjusted for age, $age^2$, sex and the interaction between them, and 10 PCs. PLINK2 (v2.00a2LM 31 March 2018 release) was used for the analysis [7]**.** Details on the sample size used for each GWAS are presented in **Supplementary Table 2** and **Supplementary Table 3**. As described in the methods we tested the above phenotypes for genetic correlation with BCC and SCC and the results are presented in **Figure 1**, **Supplementary Table 1**, **Supplementary Table 2,** and and only 22 phenotypes met the criteria to be included in the MTAG model for analysis.

**The Electronic Medical Records and Genomics Network (eMERGE ) and the KC GWAS**

eMERGE is a research biorepository (N~19,000) in the USA comprising five electric medical databases; the Group Health Cooperative Biobank (by Group Health Cooperative), Personalised Medicine Research Project (by Marshfield Clinic Research Foundation), Vascular Diseases Biorepository (by Mayo Clinic), Nugene Project (by Northwestern University) and BioVu (by Vanderbilt University) [8,9]. Detailed description on participant enrollment, genotype data processing and the study design have been published before [8–12], and further information can be accessed through the database of Genotypes and Phenotypes (dbGaP, study accession: phs000360.v3.p1). In our present study, 10,321 participants of European ancestry (1,565 KC cases and 8,756 KC free controls) were included in the KC GWAS. Cases were only considered if a participant reported a cancer code on two separate occasions when answering the survey questionnaire. Data on KC for was collected using ICD 9 codes and participants were genotyped using Illumina Human660W-Quad_v1_A array (San Diego, CA, USA) [8]. We retained SNPs with MAF > 1%.

**The Resource for Genetic Epidemiology Research on Aging (GERA) Cohort and the all-cancer GWAS**

GERA cohort is a constituent study of Kaiser Permanente Research Program on Genes, Environment, and Health (RPGEH) based in the USA. It comprises about 78,000 participants with both genotype and phenotype data called using The International Classification of Diseases, Ninth Revision, Clinical Modification (ICD-9-CM) codes. Further details on RPGEH and GERA are elaborated on the database of Genotypes and Phenotypes (dbGaP, phs000674.v3.p3). In particular, participants were genotyped using the Affymetrix Axiom arrays [13]. In our analysis cases for all-cancer were identified with ICD-9-CM codes for any cancer while controls had no history of any cancer, and data were accessed through dbGaP (study accession: phs000674.v3.p3). Cases were only considered if a participant reported a cancer code on two separate occasions when answering the survey questionnaire. After performing genotype quality procedures, we retained 61,662 self-report European ancestry participants with low genotype missingness (< 3%) and minimal ancestry divergence European ancestry (> 6 sd of PC 1 or PC2 from the HapMap phase 3 CEU population) [14]. We also retained SNPs with MAF of >1%, call rate >95% and HWE P-value > 1×10-6. PLINK 1.9 [7] was used for genotype cleaning. Next we used the Michigan Imputation Server [6] and the HRC reference panel, version r1.1 2016 [5] for imputation. SNPs with an imputation quality score > 0.3 and MAF >1% were retained for association analysis.

Next, we used SAIGE [15] to conducted a case-control GWAS on all-cancer on 61,662 participants of European ancestry (18,621 cases and 43,041 controls), and adjusted for sex and 10 PCs. The subsequent GWAS were explored for genetic correlation with BCC and for MTAG analysis as described before (**Methods**).

### Publicly accessed GWAS

#### *Cutaneous Melanoma*

We accessed and utilized summary statistics for the largest and most recently published CM GWAS meta-analysis, that included cohorts from Australia, Europe and America with participants of European descent [16]. Detailed information of the GWAS meta-analysis including; the included cohorts, quality control metrics and other statistical analyses are published elsewhere [16], and summarised here. In brief, the meta-analysis was restricted to SNPs with MAF > 0.5% and imputation quality score > 0.5. For our study, we used GWAS summary data for 81,415 participants of European ancestry from 21 cohorts that included 30,134 histo-clinically confirmed CM cases (**Supplementary Table 9**). Thus, 23andMe participants as well as the UKB self-reported CM participants were excluded.

#### *Hypothyroidism, type 1 diabetes, rheumatoid arthritis, and vitiligo*

In order to assess the auto-immune traits hypothyroidism, type 1 diabetes and rheumatoid arthritis we accessed their respective publicly available data. Details on how each GWAS was conducted are published elsewhere [15]. In summary, each GWAS was conducted using the UKB resource and SAIGE software [15] on participants of European ancestry. In addition, GWAS summary statistics on vitiligo [17] were utilised. The sample size and number of cases (where applicable) for each trait is presented in **Supplementary Table 1** and **Supplementary Table 2**.

#### *Educational attainment, smoking, and body mass index*

Using publicly available GWAS data, we also accessed and utilised summary statistics for educational attainment [18], smoking (cigarettes per day) [19], and body mass index [20]**.** Details on the sample size are summarised in **Supplementary Table 1** and **Supplementary Table 2**. All GWAS were conducted in participants of European ancestry and excluded data from 23andMe.

**SUPPLEMENTARY FIGURES**

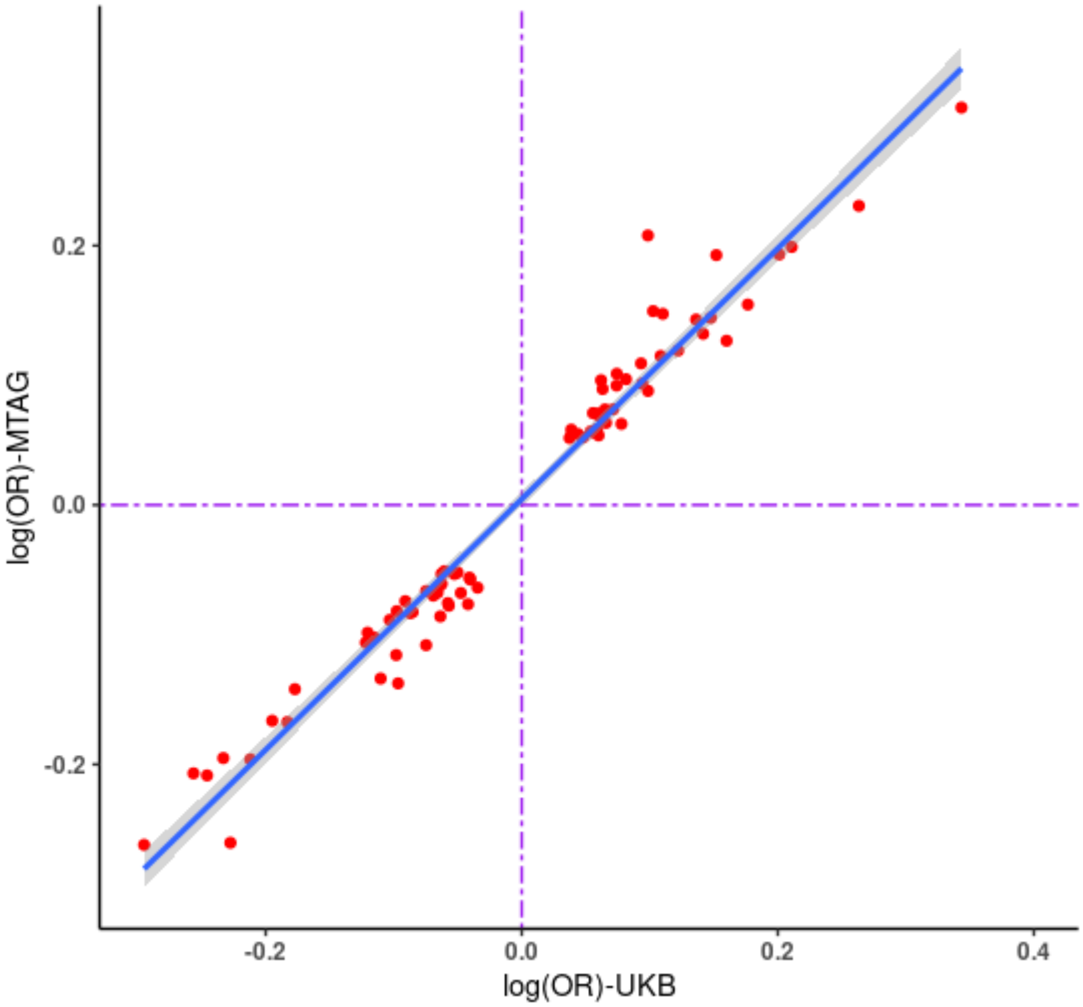


**Supplementary Figure 1:** Plot for the effect estimates (log odds ratio) for BCC MTAG versus BCC UKB GWAS excluding genomic regions with very large effect sizes including; *HLA, ASIP, IRF4, MC1R, SLC45A2*, and *CDKN2A*.

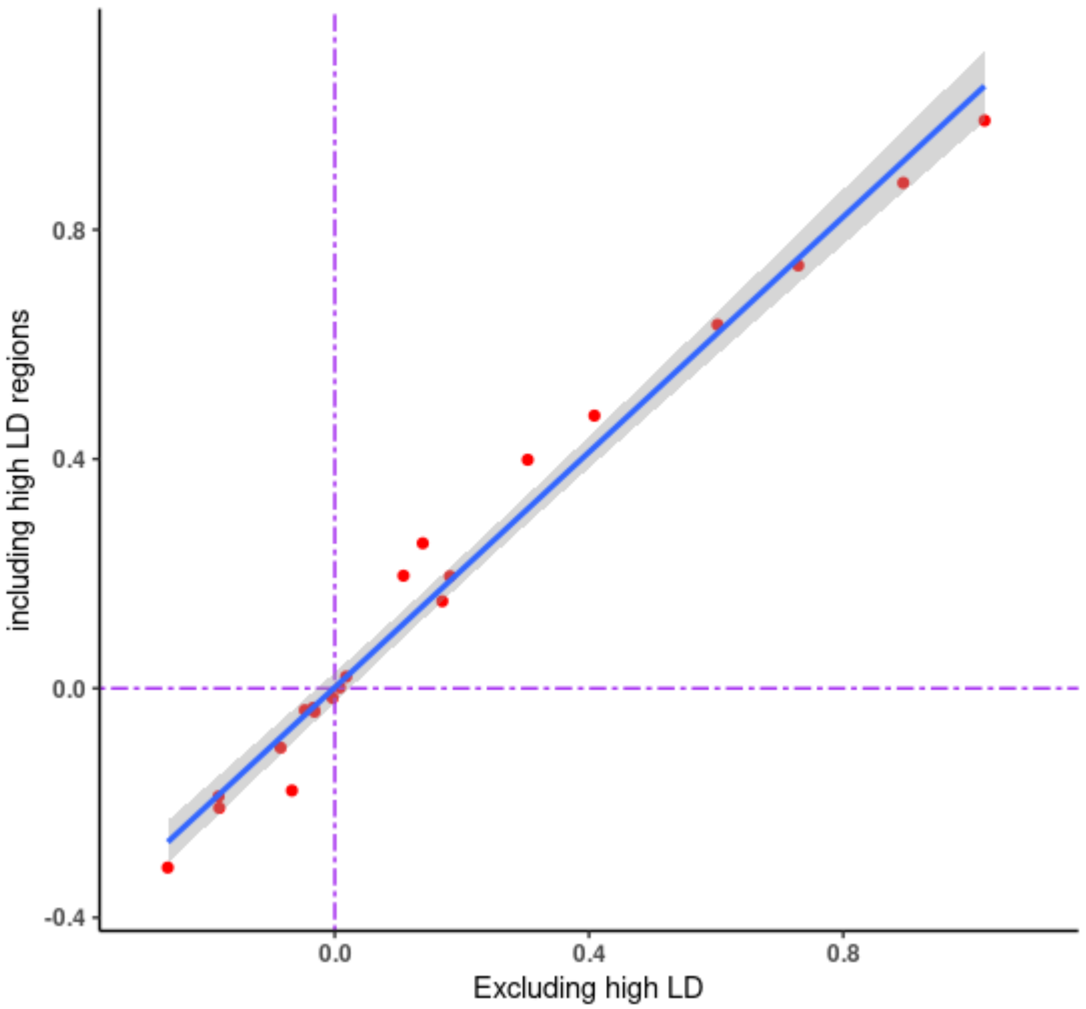


Supplementary Figure 2: Genetic correlation between BCC and 21 related traits after excluding genomic regions with very large effect sizes including; *HLA, ASIP, IRF4, MC1R, SLC45A2*, and *CDKN2A*.

### REFERENCES


1. Sudlow, C. *et al.* UK biobank: an open access resource for identifying the causes of a wide range of complex diseases of middle and old age. *PLoS Med.* **12**, e1001779 (2015).
2. Bycroft, C. *et al.* The UK Biobank resource with deep phenotyping and genomic data. *Nature* **562**, 203–209 (2018).
3. Loh, P.-R. *et al.* Efficient Bayesian mixed-model analysis increases association power in large cohorts. *Nat. Genet.* **47**, 284–290 (2015).
4. Olsen, C. M. *et al.* Cohort profile: The QSkin Sun and Health Study. *Int. J. Epidemiol.* **41**, 929–929i (2012).
5. McCarthy, S. *et al.* A reference panel of 64,976 haplotypes for genotype imputation. *Nat. Genet.* **48**, 1279–1283 (2016).
6. Das, S. *et al.* Next-generation genotype imputation service and methods. *Nat. Genet.* **48**, 1284–1287 (2016).
7. Chang, C. C. *et al.* Second-generation PLINK: rising to the challenge of larger and richer datasets. *Gigascience* **4**, 7 (2015).
8. McCarty, C. A. *et al.* The eMERGE Network: a consortium of biorepositories linked to electronic medical records data for conducting genomic studies. *BMC Med. Genomics* **4**, 13 (2011).
9. Kho, A. N. *et al.* Electronic medical records for genetic research: results of the eMERGE consortium. *Sci. Transl. Med.* **3**, 79re1 (2011).
10. McCarty, C. A., Peissig, P., Caldwell, M. D. & Wilke, R. A. The Marshfield Clinic Personalized Medicine Research Project: 2008 scientific update and lessons learned in the first 6 years. *Per. Med.* **5**, 529–542 (2008).
11. Kukull, W. A. *et al.* Dementia and Alzheimer Disease Incidence. *Archives of Neurology* vol. 59 1737 (2002).
12. Roden, D. M. *et al.* Development of a large-scale de-identified DNA biobank to enable personalized medicine. *Clin. Pharmacol. Ther.* **84**, 362–369 (2008).
13. Banda, Y. *et al.* Characterizing Race/Ethnicity and Genetic Ancestry for 100,000 Subjects in the Genetic Epidemiology Research on Adult Health and Aging (GERA) Cohort. *Genetics* **200**, 1285–1295

(2015).

14. International HapMap Consortium. The International HapMap Project. *Nature* **426**, 789–796 (2003).

15. Zhou, W. *et al.* Efficiently controlling for case-control imbalance and sample relatedness in large-scale genetic association studies. *Nat. Genet.* **50**, 1335–1341 (2018).

16. Landi, M. T. *et al.* Genome-wide association meta-analyses combining multiple risk phenotypes provide insights into the genetic architecture of cutaneous melanoma susceptibility. *Nat. Genet.* **52**, 494–504 (2020).

17. Jin, Y. *et al.* Genome-wide association studies of autoimmune vitiligo identify 23 new risk loci and highlight key pathways and regulatory variants. *Nat. Genet.* **48**, 1418–1424 (2016).

18. Okbay, A. *et al.* Genome-wide association study identifies 74 loci associated with educational attainment. *Nature* **533**, 539–542 (2016).

19. Liu, M. *et al.* Association studies of up to 1.2 million individuals yield new insights into the genetic etiology of tobacco and alcohol use. *Nat. Genet.* **51**, 237–244 (2019).

20. Yengo, L. *et al.* Meta-analysis of genome-wide association studies for height and body mass index in ∼700000 individuals of European ancestry. *Hum. Mol. Genet.* **27**, 3641–3649 (2018).
